# Fc-enhanced anti-CTLA-4 depletes tumor-infiltrating regulatory T cells to augment immune effects of androgen ablation in high-risk prostate cancer

**DOI:** 10.1101/2024.09.09.24313308

**Authors:** Casey R. Ager, Aleksandar Obradovic, Patrick McCann, Matthew Chaimowitz, Alexander L. E. Wang, Neha Shaikh, Parin Shah, Samuel S. Pan, Caroline J. Laplaca, Renu K. Virk, Jessica C. Hill, Collin Jugler, Grace DeFranco, Nilika Bhattacharya, Kade R. Copple, Howard I. Scher, Guarionex Joel DeCastro, Christopher B. Anderson, James M. McKiernan, Catherine S. Spina, Mark N. Stein, Karie Runcie, Charles G. Drake, Andrea Califano, Matthew C. Dallos

**Author notes:** **Correspondence:** Dr. Matthew Dallos. These authors contributed equally (Casey R. Ager, Aleksandar Obradovic).

## Abstract

Despite high rates of post-surgical recurrence in men with high-risk localized prostate cancer (PCa), there is currently no role for neoadjuvant therapy. Tumor infiltrating regulatory T cells (TI-Tregs) limit the antitumor effects of presurgical androgen deprivation therapy (ADT). Therefore, we designed a neoadjuvant clinical trial to test whether Treg depletion via a non-fucosylated anti-CTLA-4 antibody (BMS-986218) is feasible and augments response to ADT. In this single-center, two-arm, open-label study, 24 men with high-risk localized PCa were randomized to ADT with or without BMS-986218 prior to radical prostatectomy. Treatment was well tolerated and feasible. Mechanistic studies indicated BMS-986218 depleted TI-Tregs by engaging CD16a/FCGR3A on tumor macrophages, modulated dendritic cells (DCs), and augmented T cell priming. Depth of Treg depletion and increased DC frequencies were quantitatively associated with improved clinical outcome. Overall, this study supports the feasibility and biological activity of neoadjuvant immunotherapy with ADT + Fc-enhanced anti-CTLA-4 in high-risk localized PCa.

**Statement of Significance:** Next-generation antibodies targeting CTLA-4 have been engineered for enhanced tumor Treg depletion in patients, yet their mechanisms of action remain incompletely defined. We performed the first single cell multi-omic correlative analyses of response to a glycoengineered anti-CTLA-4 antibody and defined mechanisms associated with clinical outcome in patients with high-risk localized prostate cancer.

## Introduction

To date, immune checkpoint blockade immunotherapy (ICB) has proven largely ineffective for patients with prostate cancer (PCa)^1–3^. Agents targeting the PD-1/PD-L1 pathway alone or in combination with androgen receptor inhibitors^4–6^, chemotherapy^7^, or PARP inhibition^8^ have failed in phase III trials^9^. Similarly, CTLA-4 blockade in a chemo-naïve population^10^ or post-docetaxel in combination with bone-targeted radiation^11^ did not meet pre-specified endpoints. However, long-term follow-up indicates a potential survival advantage of ipilimumab versus placebo^11^. Furthermore, a phase II study of combination ipilimumab and nivolumab in metastatic patients pre- or post-chemotherapy showed favorable survival compared to historical controls^12^. These findings suggest that CTLA-4 blockade may possess clinical activity in PCa that warrants further investigation, particularly if concerns regarding dose-dependent toxicity can be addressed.

One hypothesis explaining the relatively low efficacy of ICB in PCa is that baseline T cell infiltration is insufficient for ICB to elicit robust T cell-mediated antitumor immunity^1^. In addition, most immunotherapy agents have first been tested in advanced disease settings where the PCa tumor microenvironment (TME) is generally more immunosuppressive, thereby potentially limiting therapeutic efficacy. Interestingly, in localized disease, standard of care androgen deprivation therapy (ADT) increases CD8 T cell infiltration into primary PCa. This finding has been replicated across multiple patient cohorts and murine models^13–15^. However, a counter-regulatory influx of immunosuppressive CD4^+^FoxP3^+^ regulatory T cells (Tregs) inhibits the inflammatory effects of ADT, implicating adaptive Treg resistance as a critical inhibitory mechanism^13,14^. As such, depletion of tumor-infiltrating Tregs (TI-Tregs) by anti-CTLA-4 antibodies with enhanced antibody dependent cellular cytotoxicity or phagocytosis (ADCC/P) extends depth and duration of response to ADT in murine models, whereas non-depleting anti-CTLA-4 antibodies or PD-1 blockade in the same setting is ineffective^13^. These Fc-enhanced agents have also been hypothesized to have an improved therapeutic index, enabling dosing strategies that deliver improved activity with reduced toxicity^16^. Therefore, we reasoned that inhibiting TI-Treg number and/or function in combination with ADT may effectively potentiate ICB efficacy in PCa.

Methods for depleting TI-Tregs have been studied thoroughly in pre-clinical models but have not been consistently validated in the clinic. Antibodies targeting cell surface proteins enriched on TI-Tregs such as CTLA-4, CD25, CCR4, TIGIT, and CCR8 have either failed to show Treg depletion activity and/or efficacy in clinical trials or remain under investigation^17–20^. We thus sought to further investigate anti-CTLA-4 in this context, given its potential clinical activity in PCa^11^ and the fact that the first-generation anti-CTLA-4 antibodies ipilimumab and tremelimumab were not optimized for Treg depletion. Tremelimumab is a human IgG2 antibody with relatively low affinity for the FcγRs such as CD16a (*FCGR3A*) that mediate ADCC/P, while ipilimumab is a human IgG1 isotype that can elicit ADCC/P. However, existing clinical data suggest ipilimumab is a weak depleter, with evidence of Treg depletion restricted to patients with a high- affinity germline polymorphic variant of *FCGR3A* (V158F)^20,21^. To overcome this limitation, antibodies targeting CTLA-4 bearing non-fucosylated Fc regions have been developed to increase binding affinity for CD16a/*FCGRIIIA*, resulting in enhanced ADCP/ADCC^22^. However, it remains unclear whether non-fucosylated anti-CTLA-4 antibodies are more effective in depleting TI-Tregs in patients. Furthermore, whether these next-generation Fc-enhanced antibodies can overcome adaptive Treg resistance following ADT is unknown.

To address this challenge, we designed a randomized neoadjuvant clinical trial to evaluate the feasibility and immunological activity of non-fucosylated ipilimumab (BMS- 986218; hereafter anti-CTLA4-NF) with ADT versus ADT alone prior to surgery in men with high-risk localized prostate cancer (NeoRED-P; NCT04301414). This window-of-opportunity trial enabled careful interrogation of the immunological mechanisms of response to anti-CTLA4-NF through orthogonal immunoprofiling by single cell RNA sequencing (scRNAseq), mass cytometry (CyTOF), and spatial proteomics (multiplex immunofluorescence). We also leveraged the novel RNA-seq-based pipeline PISCES to perform robust protein activity level assessment of tumor microenvironment subpopulations and of their drug-mediated depletion, thus bridging the transcriptomic and proteomic datasets. Pre-clinical validation studies in the syngeneic MycCaP PCa model closely mirrored observed immunological correlates of clinical response. Our findings demonstrate the feasibility of ADT plus anti-CTLA4-NF in the neoadjuvant setting and provide a first-in-human examination of whether Fc-engineered non-fucosylated anti-CTLA4 antibody variants mediate TI-Treg depletion, while identifying additional putative mechanisms by which anti-CTLA4-NF antibodies may augment antitumor immune responses in PCa.

## Results

### Safety and feasibility of neoadjuvant ADT + anti-CTLA4-NF

Between February 26, 2020, and November 15, 2022, 24 patients were enrolled at Columbia University Irving Medical Center. For correlative studies, samples from a cohort of 12 stage- and grade-matched treatment-naïve patients were collected as untreated controls. The first 4 trial patients received ADT with degarelix acetate + anti- CTLA4-NF as a safety lead-in, after which 20 patients were randomized 1:1 to ADT-only or ADT + anti-CTLA4-NF arms (Figure 1A). One patient in the ADT + anti-CTLA4-NF arm did not receive surgery due to operating room closure during the COVID-19 pandemic, and one patient in each arm was lost to follow-up for reasons unrelated to treatment (Figure 1B). Of note, one patient treated with ADT + anti-CTLA4-NF died as the result of a second unrelated malignancy while in follow-up. Baseline characteristics of the trial patients are summarized in Table 1. The arms were well balanced with respect to age, race and ethnicity, pre-treatment serum PSA or testosterone, rate of nodal involvement, and rate of surgical margin involvement. Patients in the ADT + anti- CTLA4-NF arm had a higher frequency of Gleason Grade Group 5 disease, although this difference was not statistically significant. Tumor exome and mutational profiling of all patient-derived samples showed 12 patients with AR-V7 splice variants (38%), 8 patients exhibiting TMRPSS2:ERG fusion events (25%), 4 patients bearing mutated *TP53* (13%), 3 patients with copy number loss or mutations in *PTEN* (9%), and 2 patients each with alterations *in APC* and *CTNNB1* (6%; Supplementary Figure 1A). Of note, one patient in the ADT + anti-CTLA4-NF arm was found to exhibit high tumor mutational burden (TMB-H; 26 mutations/Mb), associated with a germline mutation in *MSH2*.

**Figure 1:**
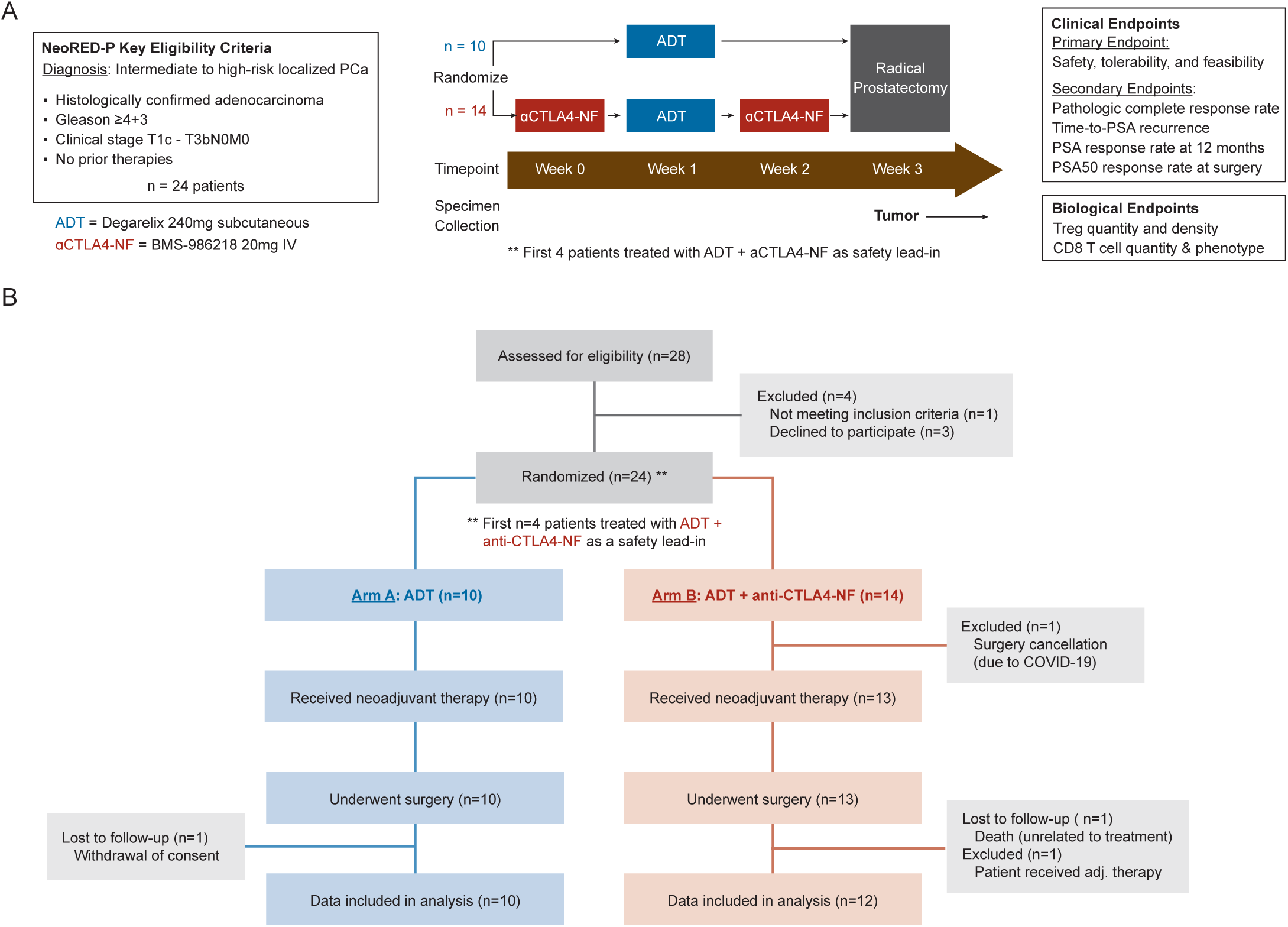
NeoRED-P trial design and consort diagram. **(A)** Schematic of NeoRED-P clinical trial design. **(B)** NeoRED-P trial consort diagram.

**Table 1.** Patient Baseline Demographics and Disease Characteristics.

| **Characteristic** | **ADT**<br>**N=10<sup>1</sup>** | **ADT + αCTLA-4-NF**<br>**N=14<sup>1</sup>** |
| --- | --- | --- |
| **Age** |  |  |
| Median (Range) | 66.0 (52.0, 72.0) | 65.5 (52.0, 77.0) |
| **Race, n (%)** |  |  |
| White | 6 (60%) | 9 (64%) |
| Black | 2 (20%) | 2 (14%) |
| Asian | 1 (10%) | 0 (0%) |
| Other | 1 (10%) | 3 (21%) |
| **Ethnicity, n (%)** |  |  |
| Not Hispanic | 7 (70%) | 12 (86%) |
| Hispanic | 3 (30%) | 2 (14%) |
| **PSA<sup>2</sup>** |  |  |
| Median (Range) | 18.44 (2.37, 94.40) | 9.08 (4.57, 364.70) |
| **Testosterone** |  |  |
| Median (Range) | 420.30 (153.90, 553.30) | 399.50 (273.80, 783.30) |
| **Grade group, n (%)** |  |  |
| 3 | 5 (50%) | 2 (14%) |
| 4 | 3 (30%) | 5 (36%) |
| 5 | 2 (20%) | 7 (50%) |
| **T stage, n (%)** |  |  |
| T2 | 5 (50%) | 4 (29%) |
| T3a | 2 (20%) | 5 (36%) |
| T3b | 3 (30%) | 5 (36%) |
| **Nodal involvement, n (%)** | 2 (20%) | 3 (21%) |
| **Surgical margin involvement, n (%)** | 3 (30%) | 3 (21%) |
| **2-year recurrence-free probability** |  |  |
| Median (Range) | 69.50 (8.00, 88.00) | 55.00 (4.00, 82.00) |
| **VHR, n (%)<sup>3</sup>** | 8 (80%) | 12 (86%) |

<sup>1</sup> Median (Range) for continuous variables; N (%) for categorical variables
<sup>2</sup> Prostate-specific antigen (PSA), ng/mL
<sup>3</sup> Very high-risk (VHR) group defined by National Comprehensive Cancer Network (NCCN) criteria as Gleason ≥8 or PSA >20 ng/ml or clinical T3 stage or higher

Treatment related adverse advents (TRAE) of any grade were observed in 80% of ADT- only patients and 71% of ADT + anti-CTLA4-NF patients and were generally attributed to ADT, with injection site reactions the most common events in both arms (Table 2).

**Table 2.** Treatment-related adverse events.

| Treatment-related adverse events (TRAEs)<sup>1</sup> | ADT<br>N = 10 |  | ADT + αCTLA-4-NF<br>N = 14 |  |
| --- | --- | --- | --- | --- |
|  | All grades | Grade ≥3 | All grades | Grade ≥3 |
| **Any treatment-related adverse event (TRAE)** | 8 (80) | — | 10 (71) | 1 (7.1) |
| **Gastrointestinal disorders** | — | — | 3 (21) | — |
| Colitis | — | — | 1 (7.1) | — |
| Diarrhea | — | — | 1 (7.1) | — |
| Elevated transaminases | — | — | 1 (7.1) | — |
| **General disorders and administration site conditions** | 7 (70) | — | 8 (57) | — |
| Chills | 1 (10) | — | — | — |
| Fatigue | — | — | 4 (29) | — |
| Hot flashes | 5 (50) | — | 6 (43) | — |
| Injection site reaction | 6 (60) | — | 7 (50) | — |
| **Investigations** | — | — | — | 1 (7.1) |
| Lipase increased | — | — | — | 1 (7.1) |
| **Musculoskeletal and connective tissue disorders** | — | — | 1 (7.1) | — |
| Bone pain | — | — | 1 (7.1) | — |
| **Nervous system disorders** | — | — | 1 (7.1) | — |
| Headache | — | — | 1 (7.1) | — |
| **Reproductive system disorders** | 2 (20) | — | — | — |
| Erectile dysfunction | 2 (20) | — | — | — |
| **Skin and subcutaneous tissue disorders** | — | — | 1 (7.1) | — |
| Pruritus | — | — | 1 (7.1) | — |

<sup>1</sup> Treatment-related adverse events (TRAEs) graded according to NCI CTCAE v5.0

Three Grade 1-2 gastrointestinal events were observed in patients receiving anti- CTLA4-NF. Grade ≥ 3 TRAEs were rare; a single patient in the ADT + anti-CTLA4-NF arm exhibited a Grade 3 asymptomatic serum lipase elevation that resolved without intervention and did not result in treatment discontinuation. No unexpected surgical complications were observed. Overall, the study met its primary endpoint of safety and feasibility, supporting a tolerable safety profile for the addition of anti-CTLA4-NF to ADT prior to radical prostatectomy in patients with high-risk localized PCa.

### Anti-CTLA4-NF counteracts ADT-induced TI-Treg expansion

This study was powered to address its key secondary biological endpoint; to test the hypothesis that anti-CTLA4-NF reduces TI-Treg density in ADT-treated PCa tumors. For this, we employed immunofluorescence staining to quantify the frequency of tumor- infiltrating CD4^+^FoxP3^+^ Tregs among all nucleated cells in whole-tissue FFPE slides obtained at prostatectomy from all evaluable study participants and matched untreated controls (see representative staining images in Figure 2A). For this analysis, we censored one patient with MSI^hi^ status given the known association between MSI^hi^ status, high TMB, and increased overall tumor T cell infiltration relative to MSI^WT^ patients^23^. In line with our previous findings, we found ADT significantly increases TI- Treg frequencies in the PCa tumor parenchyma (p=0.002; Figure 2B). Concurrent treatment with ADT and anti-CTLA4-NF resulted in a significant reduction in TI-Treg density as compared to ADT alone (p=0.012; Figure 2B). Thus, the trial met its key secondary biological endpoint of TI-Treg density reduction, supporting the hypothesis that anti-CTLA4-NF depletes TI-Tregs in PCa patients.

**Figure 2:**
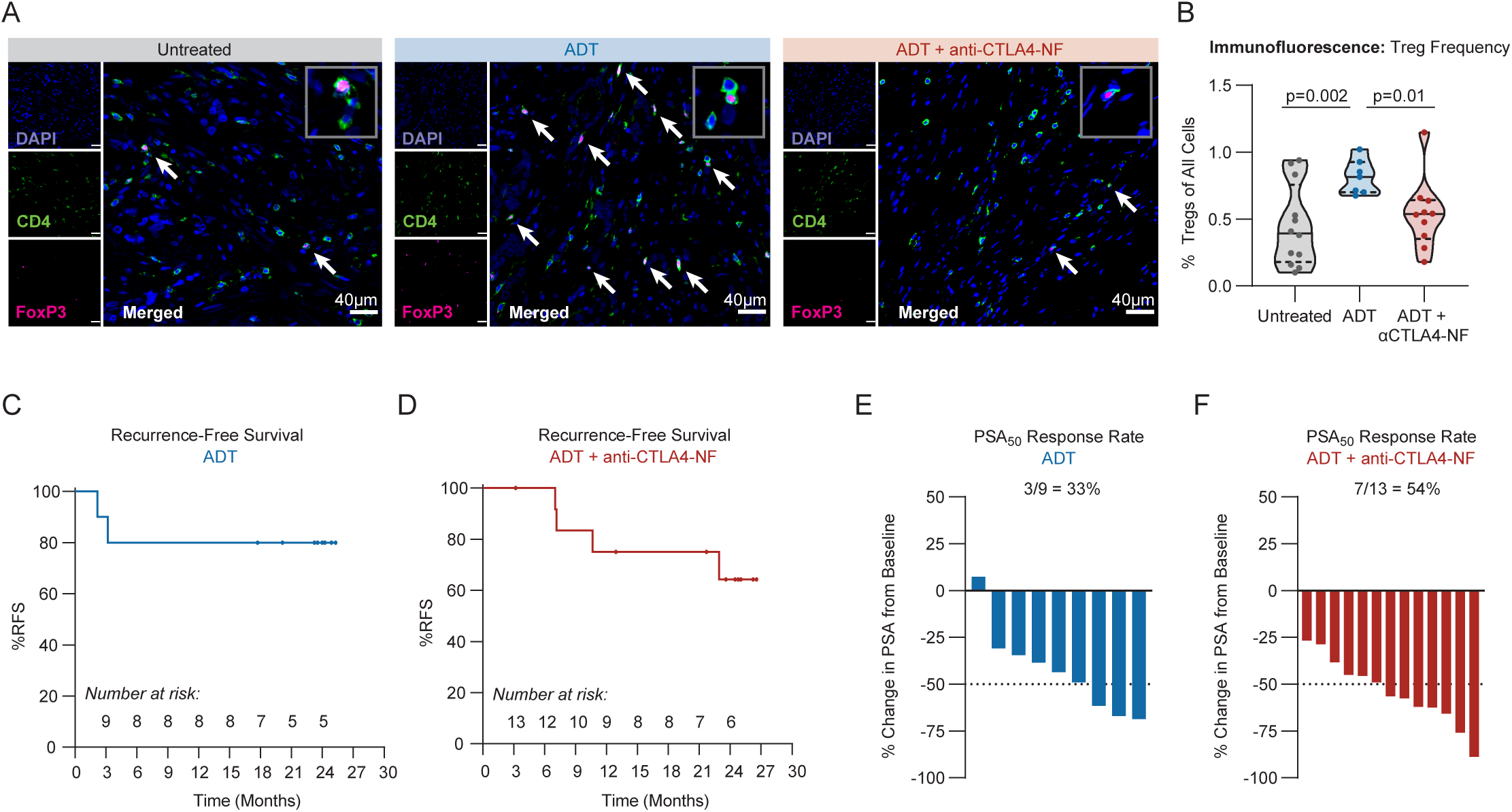
Secondary biological and clinical endpoint measures of the NeoRED-P trial. **(A)** Representative images from whole-slide immunofluorescence data for CD4 and FoxP3 with DAPI nuclear staining. Inset grey box shows a representative CD4^+^FoxP3^+^ cell at 2.5x magnification relative to the original image. Scale bar = 40μm. **(B)** Violin plot representing Treg frequency as measured by percent of DAPI^+^CD4^+^FoxP3^+^ Tregs of all DAPI^+^ cells in tumor regions iterated by treatment group in the immunofluorescence dataset. Untreated n=12; ADT n=7; ADT + anti-CTLA4-NF n=10. **(C)** Kaplan-Meier curve of PSA recurrence-free survival in patients treated with neoadjuvant ADT. **(D)** Kaplan-Meier curve of PSA recurrence-free survival in patients treated with neoadjuvant ADT + anti-CTLA4-NF. **(E)**. Percent change in serum PSA from pre-treatment to time of surgery in ADT arm patients. Dotted line denotes 50% decline in PSA (PSA50). **(F)** Percent change in serum PSA from pre-treatment to time of surgery in ADT + anti-CTLA4-NF arm patients.

This study was not powered to compare efficacy between treatment arms. However, exploratory secondary clinical endpoints of the trial included pathologic complete response rate, PSA response rate at time of surgery, and rate of undetectable PSA at 12 months. No complete pathologic responses were observed in either arm. In the ADT-only arm, 80% of patients had an undetectable PSA at 12 months, with a median recurrence-free survival of 1.61 years (IQR: 1.50–1.99 years) (Table 3). In the ADT + anti-CTLA4-NF arm, 77% had an undetectable PSA at 12 months, with a median recurrence-free survival of 1.82 years (IQR: 0.89–2.07 years) (Table 3). The overall recurrence rate in patients treated with ADT + anti-CTLA4-NF was 29% (4 of 12 evaluable patients) and 20% (2 of 10 patients) in the ADT alone arm (Table 3; Figure 2C-D). Patients in the ADT-only arm had a PSA50 response rate prior to surgery of 33%, while patients in the ADT + anti-CTLA4-NF arm had a PSA50 response rate of 54% (Table 3; Figure 2E-F). All patients recovered testosterone to baseline levels within 6 months of prostatectomy (Supplementary Figure 2). In total, the trial met its key primary and secondary study endpoints of safety, feasibility, and biological activity of anti-CTLA4-NF. This supports further clinical investigation of Fc-engineered anti-CTLA- 4 with ADT prior to surgery in high risk localized PCa.

**Table 3.** Study Outcomes.

| **Outcome** | **N** | **ADT**<br>N=10<sup>1</sup> | **ADT + αCTLA-4-NF**<br>N=14<sup>1</sup> |
| --- | --- | --- | --- |
| **PSA response rate**<sup>2</sup> | 22 | 3 (33%) | 7 (54%) |
| **Undetectable PSA at 12 months** | 23 | 8 (80%) | 10 (77%) |
| **Time recurrence-free (months)** | 23 |  |  |
| *Median (Range)* |  | 23.8 (2.2, 25.3) | 22.9 (3.2, 26.6) |
| **Overall recurrence** | 24 | 2 (20%) | 4 (29%) |

<sup>1</sup> Median (Range) for continuous variables; N (%) for categorical variables
<sup>2</sup> PSA response rate defined as >50% decrease in PSA prior to prostatectomy

### Anti-CTLA4-NF depletes TI-Tregs through myeloid Fc**γ**R engagement

To elucidate mechanisms underlying TI-Treg depletion and clinical response to anti- CTLA4-NF, we employed a single cell mutli-omic immune profiling approach via paired scRNA sequencing (using the 10X Genomics droplet-based platform) and mass cytometry (CyTOF) on independent, freshly processed prostatectomy specimens from trial patients treated with ADT + anti-CTLA4-NF, ADT alone, or no treatment (Figure 3A). Of the 36 patients assessed, 26 had scRNAseq data of sufficient quality and 25 had CyTOF data of sufficient quality to be included in further analyses (see Supplementary Figure 3A and Methods for QC criteria and Figure 3B for a representative schematic). For scRNAseq data analysis we utilized the novel PISCES pipeline incorporating unbiased Louvain-based clustering and metaVIPER (virtual inference of protein activity by enriched regulon analysis^24^) to quantitatively infer activity of approximately 6,500 regulatory proteins per cell^25^. Based on CITE-seq analysis and experimental validation, this approach has been shown to outperform traditional gene expression-based methods, wherein RNA dropout limits detection of key lineage- and function-determining transcripts and reduces cluster resolution (see Methods for more detail and prior studies^25–27^ for pipeline validation). Integrating cells from all patients, we stratified cell lineage metaclusters by PISCES, cross-validated cluster identity at the gene expression level by parallel SingleR^28^ and supervised annotation, and verified tumor cell cluster identity by aberrant copy number analysis using inferCNV^29,30^ (Supplementary Figure 3B, 4A-B). Within a T/NK lineage metacluster, we clearly resolved a Treg cluster as validated by enrichment of established Treg transcriptional signature scores, inferred activity of Treg lineage marker proteins by VIPER, and SingleR annotation (Supplementary Figure 4C-E). On the CyTOF dataset, we performed semi-supervised FlowSOM^31^ clustering followed by supervised UMAP embedding on all live CD45+ events and clearly resolved multiple immune populations expressing known lineage-defining protein markers, including a CD4+FoxP3+CD25+CD127^lo/-^ Treg cluster (Supplementary Figure 4F-I). Confirming the fidelity of our orthogonal cell clustering and annotation approaches for both datasets, we observed a high degree of correlation between immune lineage frequencies on a per- patient basis by scRNAseq versus CyTOF, and similar trends in immune lineage proportions between treatment groups between platforms (Supplementary Figure 4J-L). NK cells were a notable exception that were not significantly correlated across platforms. Though correlated, neutrophils were underrepresented in the scRNAseq data relative to CyTOF, as is common in droplet-based scRNAseq methodologies. In agreement with our IF results, both single cell platforms mirrored a reduction in TI-Treg frequency in patients receiving ADT + anti-CTLA4-NF as compared to ADT alone (Supplementary Figure 4M-N).

**Figure 3:**
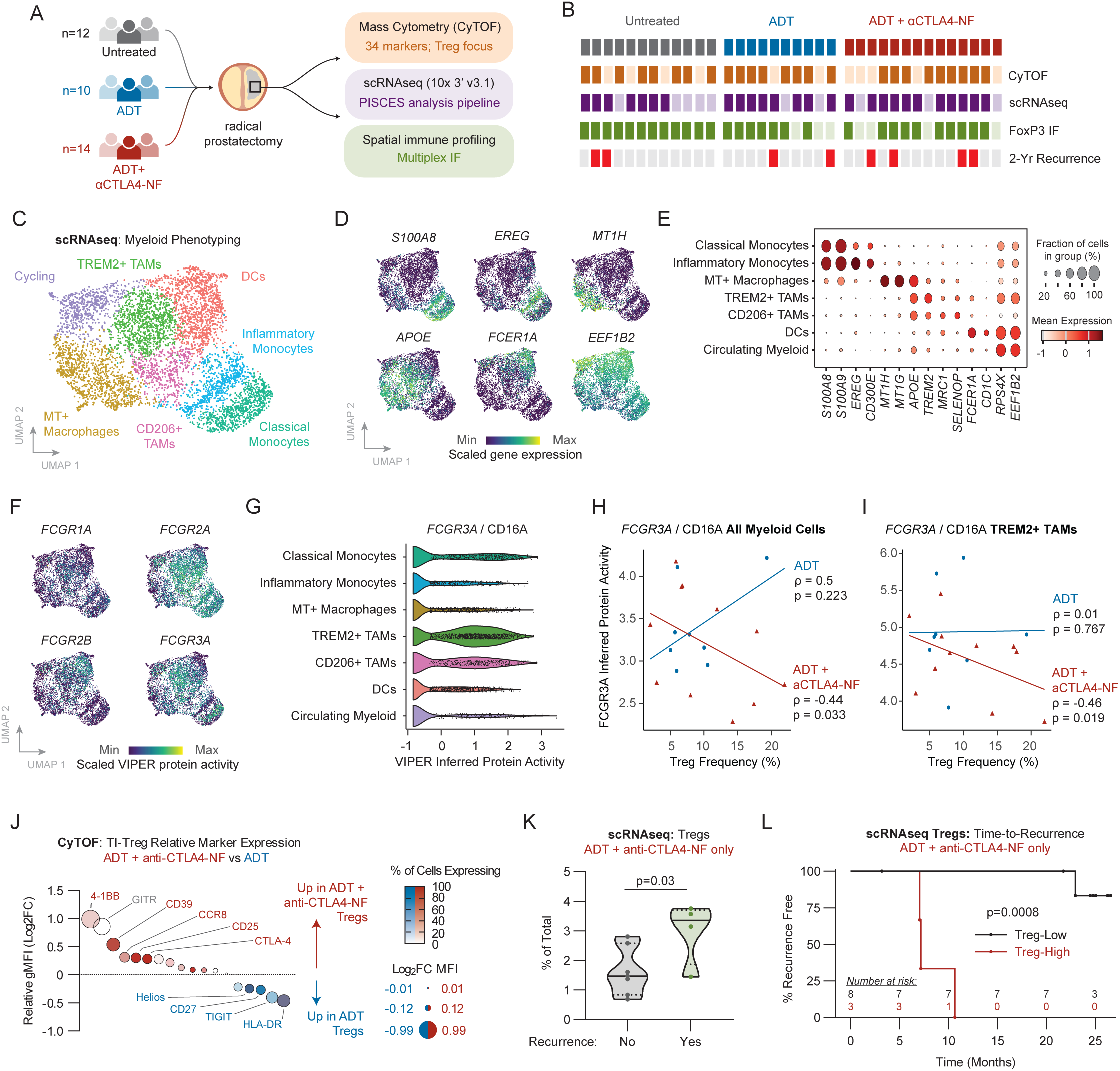
Single cell multi-omic interrogation of mechanisms underlying TI-Treg depletion and clinical response to anti-CTLA4-NF. **(A)** Schematic of immune correlates performed. **(B)** Summary of data collection and QC passing at a per-patient level for CyTOF, scRNAseq, and immunofluorescence datasets. Patients are additionally annotated according to presence or absence of PSA recurrence within 2 years post-surgery. **(C)** UMAP plot representing subclustering of myeloid cells in the scRNAseq dataset utilizing the PISCES pipeline. **(D)** Visualization of representative myeloid lineage-defining gene expression overlayed on the parent NeoRED tumor myeloid scRNAseq UMAP plot to validate cluster annotations. **(E)** Bubble plot validating expression of additional myeloid cluster defining genes. **(F)** Visualization of VIPER- inferred protein activity of select Fcγ receptors across myeloid clusters in UMAP space. **(G)** Violin plot of *FCGR3A*/CD16a inferred protein activity on a per cell level stratified by myeloid cluster. **(H)** Correlation between Treg frequency and average inferred protein activity of *FCGR3A*/CD16a across all myeloid cells on a per patient basis, stratified by treatment group. ADT n=8; ADT + anti-CTLA4-NF n=11. **(I)** Correlation between Treg frequency and average inferred protein activity of *FCGR3A*/CD16a across TREM2^+^ TAM cluster cells on a per patient basis, stratified by treatment group. Pearson correlations were utilized to evaluate statistical significance of correlations. **(J)** Bubble plot representing relative expression of phenotypic markers in Tregs from ADT + anti- CTLA4-NF group versus Tregs in the ADT only group, as calculated by log_2_ transformed ratio of average geometric MFI of each marker on Tregs from ADT + anti-CTLA4-NF versus ADT groups. Bubble size represents relative gMFI, while color intensity represents percent expression of indicated markers across all Tregs in the dataset. Red color indicates higher relative gMFI in Tregs from patients treated with ADT + anti- CTLA4-NF, while blue color indicates higher relative gMFI in patients treated with ADT only. **(K)** Violin plot representing frequency of Tregs at time of surgery in the scRNAseq dataset stratified by 2-year PSA recurrence status. No recurrence n=7; recurrence n=4. Two-tailed Welch’s t-test was performed to evaluate statistical significance. **(L)** Kaplan- Meier curve representing time-to-PSA recurrence in ADT + anti-CTLA4-NF treated patients stratified by Treg frequency in the scRNAseq data. Log-rank test was performed to evaluate statistical significance. Unless otherwise stated, two-tailed Welch’s t test was used to assess statistical significance.

In pre-clinical models, antibody-mediated Treg depletion by anti-CTLA4 requires interactions between the anti-CTLA4 Fc region and activating FcγRs on tumor- associated macrophages (TAMs)^32,33^. To determine whether macrophage FcγR expression was associated with Treg depletion efficiency in response to ADT + anti- CTLA4-NF, we first annotated macrophage populations in the scRNAseq dataset by both supervised manual annotation and through systematic testing of established PCa macrophage gene expression signatures^34^ (Figure 3C). We identified 3 putative macrophage subsets: a recently described prostate-specific MT^+^ macrophage subset^34^ and two subsets of TAMs defined by TREM2 and CD206 (*MRC1)*, respectively. Of these, TREM2^+^ and CD206^+^ TAMs exhibited the highest predicted protein activity for multiple FcγRs (Figure 3F), and TREM2^+^ TAMs showed the highest activity of the activating FcγR CD16a/*FCGR3A* (Figure 3G). To test the hypothesis that anti-CTLA4- NF depletes Tregs through CD16a/*FCGR3A* on TAMs, we examined whether CD16a/*FCGR3A* protein activity was correlated with intratumoral TI-Treg frequencies on a per-patient basis. We found predicted CD16a/*FCGR3A* protein activity across all myeloid subsets to be significantly correlated with reduced Treg frequencies in ADT + anti-CTLA4-NF patients (p=0.033; Figure 3H). In contrast, we found no significant correlation between myeloid CD16a/*FCGR3A* protein activity and TI-Treg frequencies in patients who received ADT alone (p=0.223; Figure 3H), supporting the hypothesis that CD16a/*FCGR3A* triggers myeloid directed TI-Treg ADCP upon treatment with anti- CTLA4-NF. We also found similar correlations between TI-Treg frequencies and CD16a/*FCGR3A* activity within the TREM2^+^ TAMs specifically (Figure 3I). In total, these findings support the notion that TI-Treg depletion by anti-CTLA4-NF involves engagement of tumor macrophages through the activating FcγR CD16a/*FCGR3A*.

### Depletion of phenotypically activated Tregs is associated with favorable clinical outcome

We sought to further validate and investigate the biological activity of anti-CTLA4-NF by interrogating the molecular phenotype of residual Tregs in post-treatment tumors. We utilized the CyTOF data for this analysis, as we captured an insufficient number of Tregs by scRNAseq to permit rigorous subclustering of the TI-Treg compartment. Semi-supervised FlowSOM subclustering and PaCMAP embedding of TI-Tregs in the CyTOF dataset identified nine phenotypically distinct Treg subpopulations. These were primarily resolved by differential expression of CD45RO, Ki67, granzyme B, CD39, CCR8, HLA-DR, and/or 4-1BB (Supplementary Figure 5A-C). This analysis revealed a potential phenotypic shift in residual Tregs for patients treated with anti-CTLA4-NF marked by a loss of CD39- TI-Tregs and enrichment in 4-1BB+ Tregs (Supplementary Figure 5D-E). To validate these observations, we measured the fold change in geometric MFI (gMFI) of all phenotypic markers in our CyTOF panel between Tregs in patients treated with ADT versus patients treated with anti-CTLA4-NF + ADT. We found Tregs in tumors after anti-CTLA4-NF exposure were enriched in expression of activation markers 4-1BB, CD39, CCR8, CTLA-4, and CD25, suggesting that TI-Tregs remaining after anti-CTLA4- NF have an activated phenotype (Figure 3J).

Given this observation, we next examined whether the depth of TI-Treg depletion observed in treated patients associated with better clinical outcomes. For this, we stratified patients that received ADT + anti-CTLA4-NF by recurrence (n=4) versus non-recurrence (n=7) within 2 years following surgery. We limited this analysis to the scRNAseq dataset, as we did not capture CyTOF data for all patients that recurred. We found that TI-Treg frequencies at time of surgery were significantly higher in patients that went on to recur (p=0.03; Figure 3K). In contrast, no other lymphocyte subset was significantly associated with clinical outcome (Supplementary Figure 5F). In addition, we stratified patients that received ADT + anti-CTLA4-NF by TI-Treg frequency and found that patients with high TI-Treg frequencies exhibited significantly shorter time-to-recurrence as compared to patients with low TI-Treg frequencies (p=0.0008; Figure 3L). There were insufficient recurrence events in the ADT alone arm to perform a statistical analysis of time-to-recurrence in ADT-treated patients stratified by TI-Treg frequency, though no trend was observed (Supplementary Figure 5G). In agreement with this data, prior studies show no association between Treg frequency and recurrence risk in localized PCa^35^. These findings thus suggest that the depth of TI-Treg depletion may be associated with response to ADT + anti-CTLA4-NF in patients with high-risk localized prostate cancer.

### Modulation of tumor DCs by ADT + anti-CTLA4-NF is associated with response

Given the role of myeloid-expressed CD16/*FCGR3A* in mediating Treg depletion and subsequent response to ADT + anti-CTLA4-NF, we next sought to investigate associations between myeloid population frequencies and clinical outcomes. After stratification of ADT + anti-CTLA4-NF patients by recurrence status, we observed intratumoral dendritic cell (DC) frequencies trended higher in patients that did not recur (p=0.06; Figure 4A). We then stratified patients by DC frequency at the time of surgery and found those with fewer tumor-infiltrating DCs exhibited a significantly shorter time to recurrence (p=0.043; Figure 4B). To further investigate this novel association between DC frequencies and anti-CTLA4-NF response, we returned to the MycCaP pre-clinical model of castration-sensitive PCa previously used to assess ADCC-competent versus ADCC-incompetent anti-CTLA4 antibodies in combination with ADT^13^ (Figure 4C). We implanted 8–10-week-old male FVB mice with syngeneic MycCaP tumors and performed chemical castration with degarelix acetate (ADT) when tumors averaged ∼200mm^3^ in volume. Mice received a single dose of ADT alone or in combination with three doses of anti-CTLA-4 (clone 9D9; 5 mg/kg) administered IP at three-day intervals. To confirm Treg depletion was mediated through FcγR-dependent ADCC, mice received either Fc-competent antibody (mIgG2a; anti-CTLA4 [D]) or the equivalent clone and isotype bearing a mutation that nullifies FcγR interactions^36^ (mIgG2a- LALAPG; anti-CTLA4 [ND]). These data were consistent with our prior findings that anti- CTLA4 (D), but not anti-CTLA4 (ND), augments response to ADT in the MycCaP model (Figure 4D-E). Following treatment, tumors and draining lymph nodes were harvested and profiled by 45-parameter spectral flow cytometry, and data were analyzed by semi-supervised FlowSOM clustering and supervised UMAP or unsupervised PaCMAP embedding for visualization (Figure 4F; Supplementary Figure 6A-B). ADT + anti-CTLA4 (D) significantly reduced Treg frequencies compared to the ADT and ADT + anti-CTLA4 (ND) groups, indicating robust antibody-dependent Treg depletion (p=0.0031; Figure 4G). We further found that animals treated with ADT + anti-CTLA4 (D) exhibited significantly greater frequencies of tumor-infiltrating CD11c^+^ MHC-II^hi^ DCs compared to those receiving ADT or ADT + anti-CTLA4 (ND) (p=0.0009; Figure 4H). Systematic subclustering of the tumor DC compartment revealed a striking phenotypic shift in DCs in response to anti-CTLA4 (D) (Figure 4I, Supplementary Fig 6C-D). While DC phenotypes in mice treated with ADT or ADT + anti-CTLA4 (ND) were enriched for canonical cDC1 and cDC2 markers CD103 or CD11b, respectively, nearly all DCs in mice treated with anti-CTLA-4 (D) expressed PD-L2, a marker of the DC3 subset and/or the mRegDC (mature DC enriched in immunoregulatory molecules) program (Figure 4J, Supplementary Fig 6E). The mRegDC phenotype is induced upon tumor antigen uptake, and mRegDCs are thought to possess a unique role in shaping antitumor immune responses^37^. In line with the mRegDC phenotype, these cells also upregulated CD40 (Figure 4K). Taken together, these data indicate ADCC/P competent antibodies targeting CTLA-4 modulate tumor-infiltrating DCs, and that an increased DC frequency following treatment with anti-CTLA4-NF is associated with favorable clinical outcome.

**Figure 4:**
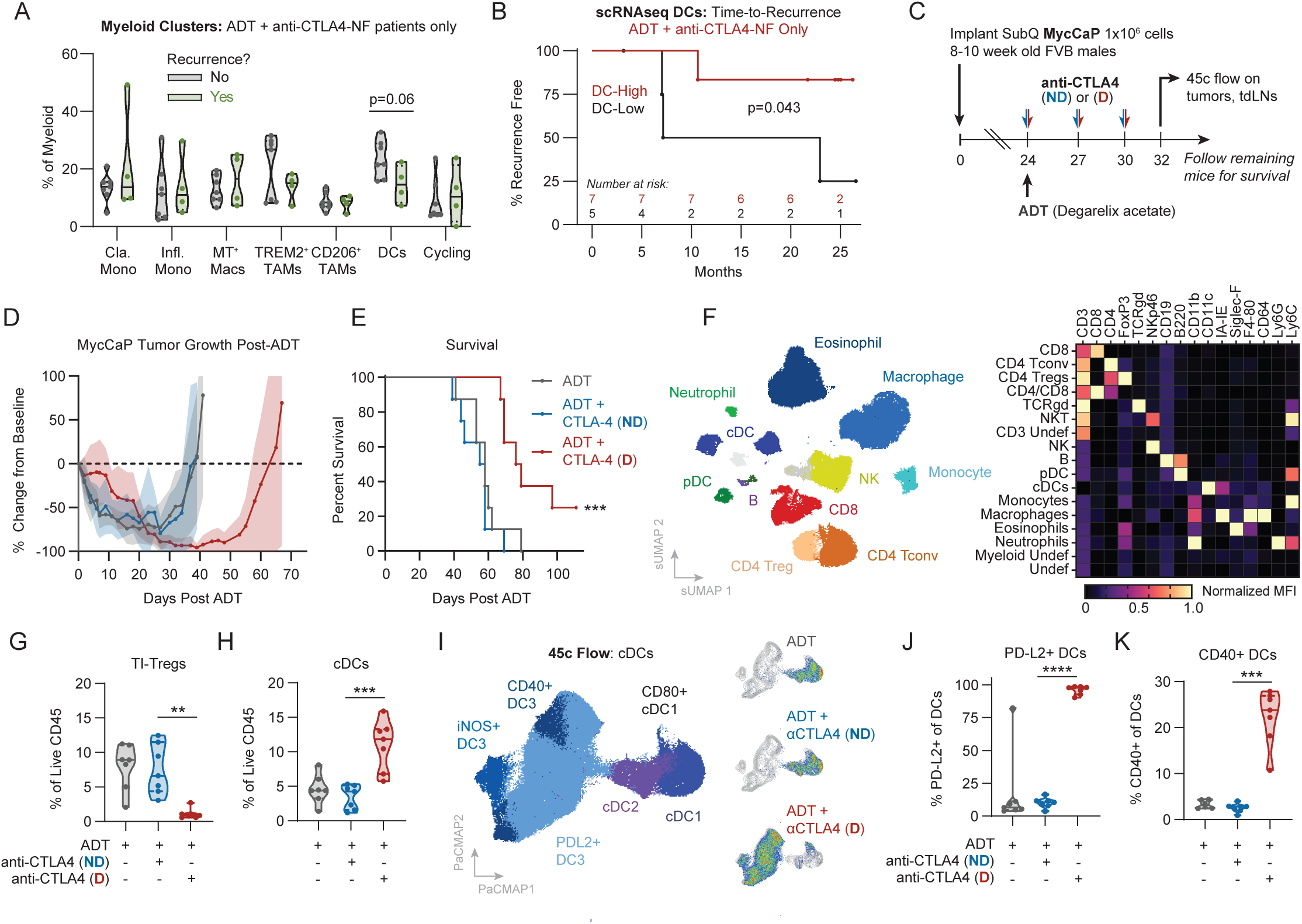
Effect of ADT + Fc-enhanced anti-CTLA-4 on DCs. **(A)** Violin plot of myeloid cluster frequencies as percent of myeloid cells stratified by patient 2-year PSA recurrence status. No recurrence n=7; recurrence n=4. Two-tailed student’s t-test was utilized to test statistical significance. **(B)** Kaplan-Meier curve representing time-to-PSA recurrence in patients stratified by dendritic cell frequency. Log-rank test was performed to evaluate statistical significance. **(C)** Schematic of pre-clinical validation experiment in the MycCaP model. **(D)** Percent change in tumor volume relative to baseline volume prior to ADT stratified by treatment group. Average values are represented by solid lines and standard deviations are represented by shaded areas. **(E)** Kaplan-Meier curve representing survival of mice shown in **(D)**. Log-rank (Mantel-Cox) test was used to evaluate statistical significance. **(F)** Left: Supervised UMAP of live CD45^+^ cells from MycCaP tumors harvested following therapy as denoted in **(D-E)**. Semi-supervised clustering was performed using FlowSOM and clusters were manually annotated according to expression of lineage-defining markers. Right: Heatmap of lineage-defining markers across all clusters. Data represents geometric MFI for each marker normalized to the minimum and maximum values across all clusters. **(G)** Frequency of TI-Tregs and **(H)** frequency of cDCs as percent of all live CD45^+^ cells, stratified by treatment group. **(I)** Left: PaCMAP representing FlowSOM-derived cDC clusters. Clusters were manually annotated according to expression of canonical marker proteins. Right: Pseudocolor representation of cDC phenotypes in PaCMAP space stratified by treatment group. **(J)** Violin plot representing frequency of PD-L2^+^ DCs as percent of all DCs stratified by treatment group. **(K)** Violin plot representing frequency of CD40^+^ DCs as percent of all DCs stratified by treatment group. Welch’s t-test was used to assess statistical significance. Murine data shown is n=8 mice per group and representative of two independent experiments each for survival and immune profiling studies.

### ADT + anti-CTLA-4-NF enhances antitumor T cell priming

Given the observed effects of anti-CTLA4-NF therapy on tumor Treg frequencies and myeloid phenotypes, we next sought to determine the effect of ADT + anti-CTLA4-NF on CD8 T cell responses in the tumor. For enhanced resolution of CD8 phenotypes in the scRNAseq data, we first refined the previously identified CD8-1 and CD8-2 clusters, removing a contaminating cell population presenting low *Cd8a* expression, moderate *Cd4* expression, and an otherwise naïve gene signature (Supplementary Figure 7A-B). Subcluster analysis of the curated CD8 T cells resulted in an optimal three cluster solution (Supplementary Figure 7C). Utilizing established pan-cancer CD8 T cell gene signatures from Zheng *et al.*^38^ and prostate CD8 T cell signatures from Tuong *et al.*^34^ we annotated these clusters as central memory (IL7R^+^ memory signature enrichment, prostate CD8 memory T cell signature enrichment, expression of CCR7), effector memory (enrichment of GZMK^+^ early TEM signature), and effector/resident memory CD8 T cells (enrichment of prostate CD8 effector T cell signature, expression of CD103, GzmB, TOX2; Supplementary Figure 7D). These subsets were not significantly altered by ADT or ADT + anti-CTLA4-NF (Supplementary Figure 7E). To further interrogate tumor-infiltrating CD8 T cell phenotypes at the protein level by CyTOF, we used semi-supervised FlowSOM clustering to define eight CD8 T cell subclusters (Figure 5A).

**Figure 5:**
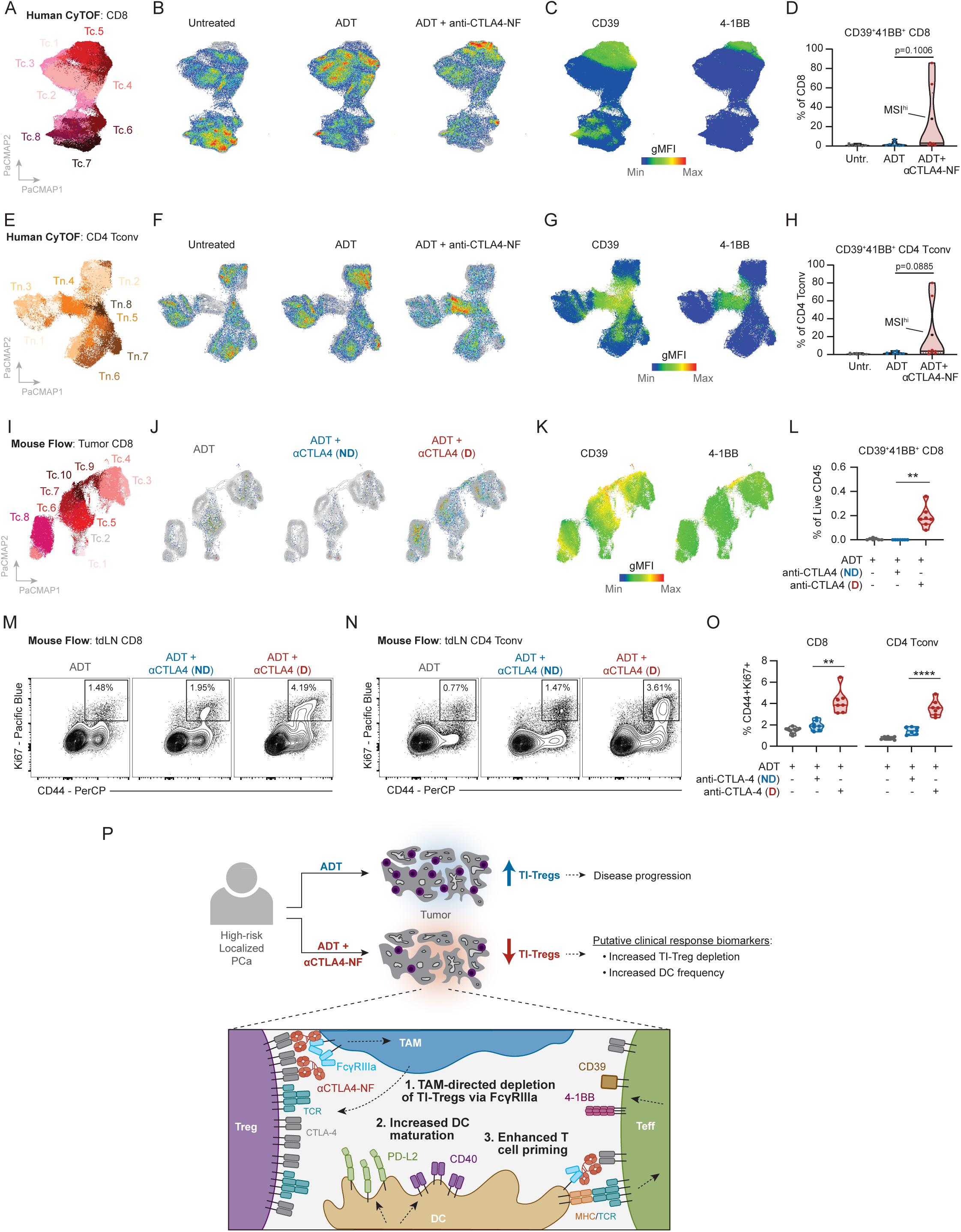
Phenotypic modulation of T cells and enhanced priming by anti-CTLA4- NF. **(A)** PaCMAP plot of NeoRED-P patient tumor-infiltrating CD8 T cells by CyTOF. Clusters were derived from FlowSOM. **(B)** Pseudocolor density plots of CD8 T cells in PaCMAP space stratified by treatment group. **(C)** Expression of CD39 and 4-1BB by geometric MFI on CD8 T cells as represented by color mapping on PaCMAP plot. **(D)** Violin plot representing frequency of manually gated CD39^+^4-1BB^+^ CD8 T cells as a percentage of all CD8 T cells stratified by treatment group. Untreated n=7; ADT n=8; ADT + anti-CTLA4-NF n=8. Single patient with MSI^hi^ status called out in plot. **(E)** PaCMAP plot of NeoRED-P patient tumor-infiltrating CD4^+^ FoxP3^-^ Tconv cells by CyTOF. Clusters were derived from FlowSOM. **(F)** Pseudocolor density plots of CD4 Tconv cells in PaCMAP space stratified by treatment group. **(G)** Expression of CD39 and 4-1BB by geometric MFI on CD4 Tconv as represented by color mapping on PaCMAP plot. **(H)** Violin plot representing frequency of manually gated CD39^+^4-1BB^+^ CD4 Tconv cells as a percentage of all CD8 T cells stratified by treatment group. Untreated n=7; ADT n=8; ADT + anti-CTLA4-NF n=8. Single patient with MSI^hi^ status called out in plot. **(I)** PaCMAP plot of MycCaP-infiltrating CD8 T cells by 45-parameter flow cytometry in response to ADT, ADT + anti-CTLA4 (ND), or ADT + anti-CTLA4 (D). Clusters were derived from FlowSOM. **(J)** Pseudocolor density plots of CD8 T cells in PaCMAP space stratified by treatment group. **(K)** Expression of CD39 and 4-1BB by geometric MFI on CD8 T cells as represented by color mapping on PaCMAP plot. **(L)** Violin plot representing frequency of manually gated CD39^+^4-1BB^+^ CD8 T cells as a percentage of all CD8 T cells stratified by treatment group. **(M)** Biaxial plots representing expression of CD44 and Ki67 on CD8 T cells in tumor draining lymph nodes of mice shown in **(I-L)**. **(N)** Biaxial plots representing expression of CD44 and Ki67 on CD4^+^FoxP3^-^ Tconv cells in tumor draining lymph nodes of mice shown in **(L-O)**. **(O)** Violin plots representing frequencies of CD44^+^Ki67^+^ CD8 and CD4 Tconv cells as a percentage of parent populations stratified by treatment group. Two-tailed Welch’s t-test was used to assess statistical significance. All murine data shown is n=7 per group and representative of two independent experiments each for survival and immune profiling studies. **(P)** Schematic overview of working model based on findings from this study.

While these clusters did not map precisely onto the three cluster solution in the scRNAseq data, we did observe clusters that most closely matched the effector memory (Tc.4; CD45RA^-^CD45RO^+^CD27^+^HLA-DR^+^PD-1^mid^) phenotype and cells that matched the effector/resident memory (Tc.6; CD45RO^-^HLA-DR^+^CTLA-4^+^TIGIT^+^GzmB^+^) phenotype and mirrored non-significant trends in the scRNAseq data after anti-CTLA4- NF (Supplementary Fig 7F-G). Notably, we observed a small cluster with clear enrichment in a subset of patients receiving ADT + anti-CTLA4-NF defined by 4-1BB and CD39 (Figure 5B-D), markers that are associated with recently primed and tumor-specific T cells^39,40^. We found the dual 4-1BB^+^CD39^+^ phenotype to also define a distinct subset of CD4^+^FoxP3^-^ conventional CD4 T cells (Tconv) that were uniquely present in some patients receiving ADT + anti-CTLA4-NF (Figure 5E-H, Supplementary Figure 7H- I). These data suggest anti-CTLA4-NF may enhance priming of tumor-specific T cell populations, either in concert with or independent of Treg depletion. To test whether this effect requires FcγR interactions, we returned to our murine model where we profiled effects of anti-CTLA4 (D) versus anti-CTLA4 (ND) in combination with ADT. We observed a strikingly similar cluster of 4-1BB^+^CD39^+^ CD8 T cells in Myc-CaP tumors that were uniquely induced by ADT + anti-CTLA4 (D) and were absent in the ADT-only and ADT + anti-CTLA4 (ND) groups (Figure 5I-L, Supplementary Fig 6F-H), indicating FcγR engagement is required for induction of the 4-1BB^+^CD39^+^ CD8 T cell phenotype following anti-CTLA4 therapy. Supporting our hypothesis that these cells represent a recently primed population, we found a significant increase in CD8 and CD4 Tconv cells bearing phenotypic markers of recent priming (CD44^+^Ki67^+^) in tumor draining LNs (tdLNs) in mice receiving anti-CTLA4 (D) relative to anti-CTLA4 (ND) or ADT alone (Figure 5M-O, Supplementary Fig 6I). This was not associated with Treg depletion in the tdLNs, but rather an increase in Treg frequency in the tdLNs as has been previously shown in the context of anti-CTLA4 therapeutics^41^ (p<0.0001; Supplementary Figure 6J). These findings support an additional putative mechanism of action of Fc engineered anti-CTLA4-NF antibodies, potentially distinct from Treg depletion, involving enhanced priming of tumor-specific CD8 and/or non-Treg CD4 Tconv cells (working model summarized in Figure 5P).

## Discussion

This study evaluated a novel next-generation non-fucosylated anti-CTLA4 antibody (BMS-986218) engineered to enhance ADCC in combination with ADT versus ADT alone prior to surgery in men with high-risk localized PCa. While most early-phase clinical studies test novel investigational agents in highly advanced, late-stage disease, our study was unique in evaluating anti-CTLA4-NF in patients with localized PCa based on prior evidence suggesting this setting would be more amenable to the distinct immunological effects of combined ADT + anti-CTLA4-NF. We demonstrated that anti- CTLA4-NF has promising biological activity in patients, consistent with its proposed mechanism of action of TI-Treg depletion and enhances T cell priming in PCa. Anti- CTLA4-NF + ADT treatment was feasible with a tolerable safety profile in the pre-surgical setting, meeting our pre-specified primary study endpoints.

To determine mechanisms underlying the immunological effects of anti-CTLA4- NF, we utilized multiple analytic methods to interrogate changes in immune phenotypes in the TME at the single-cell level. These studies revealed both expected and novel associations with treatment response. We showed for the first time in patients that a glycoengineered anti-CTLA-4 antibody can mediate TI-Treg depletion through engagement of the activating FcγR CD16a/*FCGR3A* on tumor macrophages and that the depth of Treg depletion correlates with clinical outcome. Our findings thus support the mechanism established in pre-clinical models, but for which limited clinical evidence currently exists^20,32^. These findings also support the hypothesis that first-generation anti- CTLA4 antibodies ipilimumab and tremelimumab fail to deplete TI-Tregs in humans due to inefficient engagement of CD16a on TAMs. A possible explanation for this phenomenon is that non-engineered anti-CTLA4 IgG1 antibodies preferentially engage the inhibitory FcγR *FCGR2B*/CD32b on myeloid cells, preventing TI-Treg-directed ADCC/P^42^. Complicating this mechanism is our observation that inhibitory *FCGR2B* and activating *FCGR3A* receptors are highly co-expressed on tumor associated macrophages. Additional studies are warranted to understand how co-expression patterns of *FCGR3A* and *FCGR2B* on TAMs influence efficiency of Treg depletion by Fc-engineered antibodies targeting CTLA-4. An alternative mechanism supported by our data is that tumors with low baseline frequencies of *FCGR3A*/CD16a+ TAMs – or physical segregation of these macrophages from TI-Tregs – may fail to respond to anti- CTLA4-NF. A positive role for *FCGR3A*/CD16a^+^ TAMs in the context of anti-CTLA4-NF is notable, as *FCGR3A*/CD16a^+^ TAMs are generally enriched for the immunosuppressive, M2-like phenotype (e.g. TREM2^+^ and CD206^+^ TAMs in our study) that plays a negative prognostic role in many cancers^26^. However, the role of these TAMs may vary in distinct therapeutic contexts. Recent data suggest CD206^+^ M2-like TAMs may in fact be necessary to organize antitumor immune responses^43^, which together with our findings challenges the narrative that M2-like TAMs are universally antagonistic to cancer immunotherapies. In either case, pre-clinical studies suggest anti-CTLA4-mediated FcγR engagement on tumor macrophages can elicit pro-inflammatory macrophage skewing^44^. Therefore, Fc-engineered anti-CTLA4 antibodies may deliver a broadly applicable, and possibly synergistic, dual benefit of TI-Treg depletion and M2-to-M1 macrophage skewing that may be effective in overcoming the immunosuppressive microenvironment of many tumor types that do not respond to ICB.

Although anti-CTLA4-NF exhibited TI-Treg depletion in patients with high macrophage FcγR expression, this TI-Treg depletion was incomplete. Residual Tregs in tumors following treatment with anti-CTLA4-NF appear phenotypically activated. While our current studies were not able to confirm whether these Tregs were functionally immunosuppressive, these findings indicate a potential limitation of TI-Treg depleting therapies that target a single coinhibitory receptor such as CTLA-4. We hypothesize that TI-Tregs undergoing blockade of CTLA-4 without experiencing ADCC/P (whether through engagement of *FCGR2B* on proximal TAMs or spatial segregation from ADCC/P effectors) become activated as a result of reduced coinhibitory receptor activity, as has been observed in mice receiving anti-CTLA4 or in context of anti-PD1 blockade^41,45^. Notably, we found these Tregs to be enriched in other potentially targetable surface proteins, such as CCR8, CD25, CD39, and 4-1BB. Fc competent and glycoengineered antibodies targeting CCR8 are currently in clinical development^46,47^.

Thus, co-delivery of glycoengineered antibodies targeting CTLA-4 and CCR8 may be complementary, given the potential capacity for anti-CCR8 to deplete TI-Tregs remaining following anti-CTLA4, and that anti-CTLA4 elicits enhanced T cell priming and myeloid activation. Recent studies also suggest CD4 Tconv in tumors depleted of TI-Tregs upregulate CCR8 and become functionally immunosuppressive^48^. These studies warrant additional basic and clinical studies of glycoengineered antibodies targeting CTLA4 and CCR8 in prostate and other cancers.

In addition to Treg depletion, we found that our glycoengineered anti-CTLA4 antibody enhanced T cell priming, potentially by increasing interactions between CTLA4+ effector T cells and FcγR+ dendritic cells. High tumor DC frequencies in our data were significantly associated with lack of disease recurrence in patients receiving ADT + anti-CTLA4-NF. In addition, only patients treated with anti-CTLA4-NF elicited the distinct CD39^+^41BB^+^ phenotype in tumor-infiltrating CD8 and CD4 T cells, mirroring established phenotypes for tumor-specific T cells (CD39^+^CD103^+39^) and recently primed tumor-specific cells (41BB^+^). Our pre-clinical studies in the murine syngeneic MycCaP model were in striking agreement with these clinical data, and prior *in vitro* work with human T cells and DCs in context of bacterial superantigen-mediated stimulation suggest FcγR engagement on APCs is required for enhanced priming activity of antibodies targeting CTLA-4, independent of Treg depletion^49^. Our pre-clinical findings that FcγR-engaging – but not FcγR-silenced – anti-CTLA-4 antibodies elicit a significant phenotypic shift in tumor DCs suggests a bi-directional effect on both T cells and DCs, reminiscent of observed inflammatory polarization of macrophages post-anti-CTLA-4 in other murine models^44^. Nevertheless, increasing evidence indicates spatial co- localization of effector CD8 and CD4 T cells with DCs in tumors (termed immune “triads,” “hubs” or “networks”^50,51^) is critical for productive antitumor immune responses. Further investigation is warranted to determine whether Fc-enhanced antibodies targeting CTLA-4 or other immune checkpoints can facilitate accumulation of these specialized immune clusters in human tumors, and whether this is an essential underlying mechanism of action of these therapeutics.

Our study had several limitations. One key limitation was the relatively small sample size, making interpretation of the clinical activity of ADT + anti-CTLA4-NF challenging. Indeed, our study was not designed with sufficient power to compare outcome differences between the treatment arms. In addition, our orthogonal multi-omic measures of TI-Treg depletion relied on post-treatment frequency or density comparisons between treatment groups, as opposed to matched pre- and post- treatment tissue, as freshly processed pre-treatment samples were not available. It remains possible patients in the ADT + anti-CTLA4-NF arm possessed higher baseline TI-Treg densities than untreated or ADT-treated patients, given their relative enrichment for Gleason Grade Group 5 disease. Thus the observed TI-Treg depletion activity in our dataset would be underestimated. However, comparisons between pre-treatment biopsies and surgical resection tissue may be confounded when assessing the immune TME due to inflammatory effects of biopsy collection or to spatial intratumoral heterogeneity^52^. Clinically, it is notable that development of BMS-986218 has been discontinued (NCT03110107). Nevertheless, several related second-generation depleting anti-CTLA4 antibody agents (botensilimab/AGEN1181, ONC-392, ADG216) are in clinical development. Of note, recent findings from Chand *et al.* indicate botensilimab, an Fc-enhanced DLE-mutated anti-CTLA4-hIgG1 antibody, depletes TI- Tregs in an FcγR-dependent fashion across cancer types and potentially modulates DCs, closely mirroring our pre-clinical and clinical findings with BMS-986218 in PCa^16^.

In conclusion, we demonstrate the safety and feasibility of anti-CTLA4-NF (BMS- 986218) in combination with ADT prior to surgery in men with high-risk localized prostate cancer. Our robust single cell immune profiling studies provide first-in-human mechanistic insight into macrophage-mediated TI-Treg depletion by glycoengineered antibodies targeting CTLA-4. In combination with pre-clinical modeling, these findings support both canonical and novel mechanisms underlying activity of Fc-enhanced anti- CTLA4. Our study identifies immunological correlates of clinical response to ADT + anti-CTLA4-NF to inform future studies. Given the limited efficacy and significant toxicity associated with neoadjuvant ipilimumab in prior studies involving PCa patients^53^ (also see NCT02020070), our findings provide a novel avenue to explore Fc-enhanced CTLA-4 targeted immunotherapy in patients with PCa.

## Methods

### Patients

Key eligibility criteria included histologically confirmed adenocarcinoma of the prostate (clinical stage T1c-T3b, N0, M0) with no detectable metastatic involvement of lymph nodes, bone, or visceral organs and high-risk disease as defined by central pathologic review of at least two biopsy cores with Gleason sums of ≥4+3. Patients were required to demonstrate adequate bone marrow, hepatic, and renal function. Prior exposure to radiation therapy, chemotherapy, immunotherapy, biologic therapy, or concomitant treatment with hormonal therapy and/or systemic corticosteroids (with the exception of short courses <5 days 7 days or more prior to treatment initiation) was not allowed.

Subjects were recruited at Columbia University Medical Center. See full trial protocol in Supplementary Information for additional detail.

### Clinical Trial Regulatory Oversight and Approval

This trial was approved and all amendments reviewed by the Columbia University Institutional Review Board (IRB) under the protocol number AAAS3560. The trial was registered with ClinicalTrials.gov on March 6, 2020, with the ClinicalTrials.gov ID NCT04301414. All patients provided informed consent prior to enrollment in the study.

### Clinical Trial Design

This was a single-center, randomized, two-arm, open label pilot study conducted at Columbia University Irving Medical Center to evaluate the safety, feasibility, and immunogenicity of neoadjuvant degarelix (ADT) or BMS-986218 (anti-CTLA4-NF) plus ADT prior to radical prostatectomy in men with high-risk localized prostate cancer.24 patients were enrolled in the study. The first 4 patients were assigned to receive ADT + anti-CTLA4-NF as a safety lead-in. Thereafter, 10 patients each were randomized by the study statistician to the two treatment arms. In the ADT-only arm, men received 240 mg degarelix acetate subcutaneously two weeks prior to surgery. In the ADT + anti- CTLA4-NF arm, men received 20 mg BMS-986218 intravenously on day 1 and day 15, and 240 mg degarelix acetate subcutaneously on day 8, starting 3 weeks prior to prostatectomy (Figure 1A). The clinical trial sample size was driven by detecting a pharmacodynamic effect of the agent based on the key secondary endpoint of Treg infiltration. A biologically meaningful treatment effect was deemed to be a 50% decrease in Treg infiltration with BMS-986218 plus degarelix versus degarelix alone. Previous studies show that Treg density after degarelix is 59 cells /mm2 (95% CI: 34- 85). A sample size of 12 evaluable subjects per group achieves greater than 90% power to detect a minimum difference of 29 cells /mm^2^ based on two-sided two-sample t-test with a significance level (alpha) of 0.05.

### Clinical Trial Endpoints and Assessments

The primary objective was to characterize the safety, tolerability, and feasibility of ADT with or without anti-CTLA4-NF in the neoadjuvant setting. The secondary objectives included: (i) evaluation an immune response consistent with the proposed mechanism of action of anti-CTLA4-NF, specifically the effect on intratumoral Treg density and CD8:Treg ratio, (ii) estimation of the pathologic complete response rate following neoadjuvant anti-CTLA4-NF plus ADT as compared to ADT alone, (iii) estimation of the PSA50 response rate and rate of undetectable PSA at 12 months following neoadjuvant anti-CTLA4-NF plus ADT as compared to ADT alone, and (iv) estimation of the rate of post-surgical PSA recurrence within two years of completing therapy between patients receiving ADT alone versus ADT plus anti-CTLA4-NF.

Serum PSA and testosterone levels were measured on all patients prior to therapy initiation and at time of surgery. Thereafter, serum PSA and testosterone was measured on all patients every 3 (±1) months during the first post-operative year and every 6 (±2) months during year 2 and 3 of follow up. Adverse events were monitored during the course of therapy and until at least two years post-operatively and registered according to CTCAE version 5.0. Adverse events were deemed associated or not associated with ADT and/or anti-CTLA4-NF according to the treating physician. Tumor samples from prostatectomy specimens were collected at time of surgery; fresh tissue was collected and processed as described below, and archival formal-fixed paraffin-embedded (FFPE) tissues were obtained for indicated correlative studies.

### Tumor Sample Collection & Processing

Following radical prostatectomy, surgical tissues were grossed and sectioned by pathologists at Columbia University Irving Medical Center. Up to 1 gram of tumor tissue was collected in Miltenyi MACS Tissue Storage Solution on wet ice, massed, washed with PBS, then diced with microdissection scissors to 1-2mm pieces in a 5mL Eppendorf tube. Tissue pieces were transferred into a Miltenyi C tube (0.5g per tube) in serum free RPMI, then enzymes H, R, and A were added according to the Miltenyi Human Tumor Dissociation Kit protocol. Tumors were digested on a gentleMACS Octodissociator under program 37_multi_A_01. Once finished, RPMI supplemented with 5% FBS (R5 media) was added to quench digest enzymes, then suspensions were filtered (70um) into a 15mL conical tube and spun at 400xg for 5 minutes at 4° C. Pellets were RBC lysed by resuspension in Miltenyi 1x ACK buffer for 5 minutes at RT, followed by centrifugation at 300xg for 5 min at 4° C. Pellets were washed with R5, filtered, and pelleted. Cell numbers were then quantified on a Countess II automated cell counter, then samples were split for downstream analysis by CyTOF and single cell RNA sequencing.

### Tumor Exome Sequencing

Baseline germline mutation testing was performed at trial screening using the Invitae Common Hereditary Cancers Panel (48 genes) or the Invitae Multi-Cancer Panel (70 genes). Archival formalin-fixed, paraffin-embedded (FFPE) tumor specimens with >30- 40% tumor content were sectioned at 5uM by the Herbert Irving Comprehensive Cancer Center’s (HICCC) Molecular Pathology Shared Resource (MPSR). Hematoxylin and eosin (H&E) slides were reviewed and tumor area marked by a board-certified pathologist. Tissue sections were submitted for mutational profiling to Columbia University Irving Medical Center’s Laboratory of Personalized Genomic Medicine, a CAP-accredited and CLIA-certified clinical genomics laboratory. Tissue was prepared for sequencing as previously described^54^.

Targeted next-generation sequencing of tumor DNA and cDNA was performed using the Columbia Combined Cancer Panel (CCCP), a New York State Department of Health-certified panel, covering 467 cancer-related genes Raw sequencing data were mapped to the human reference genome (GRCh37, hg19), and analyzed for single nucleotide variants, small insertions and deletions, copy number alterations, gene rearrangements and fusions, tumor mutation burden, and microsatellite instability using validated clinical pipelines. Identified genomic alterations were reviewed and interpreted for pathogenicity and clinical significance by a board-certified molecular pathologist. See supplementary Information for genes covered in all genomic testing panels and panel-specific methodologies.

### Mass Cytometry

Cell pellets were resuspended in R5 supplemented with Rh103 viability stain (1:500) in a 1.5mL protein lo-bind tube and incubated 15 min at 37° C. Cells were pelleted, washed once with R5 media, then resuspended in 49ul Cell Staining Buffer (CSB; Fluidigm) followed by addition of 1ul Fc Block (TruStain FcX; BioLegend). Samples were mixed and incubated for 5-10 minutes on ice. Separate surface and intracellular antibody cocktails were pre-mixed and stored at -80° C; during the Fc Block incubation cocktails were thawed on ice, the surface mix was spun through a PVDF centrifuge filter for 3 min at 1200rpm, then 50ul surface mix was added to the cell suspension for a final volume of 100ul. Surface staining was conducted on ice for 30 minutes, then cells were washed twice with 1mL CSB. Cells were fixed by resuspending in 500ul FoxP3 Fixation/Permeabilization buffer (eBioscience) and incubating at RT for 30 minutes.

Fixative was washed out with two successive washing steps with 1mL 1X Perm Wash. For intracellular staining, cells were resuspended in 49ul 1X PW + 1ul heparin on ice. Intracellular cocktails were diluted to 50ul with 1X PW and were spun through a PVDF membrane prior to addition to the cell solution for a final staining volume of 100ul. Cell staining was conducted on ice for 30 minutes, followed by two successive washes with 1mL 1X PW. Fully stained cells were fixed in 400ul FixIR solution (2.4% PFA + 0.25 nM Ir-191/193) at room temperature for 30 minutes. Samples were washed with 600ul CSB and spun for 4 min at 600 xg, and washed again with 1mL CSB. Samples were either resuspended in CSB and held at 4° C for a maximum of 72 hours before being acquired on a CyTOF Helios at the Columbia University Human Immune Monitoring Core (HIMC), or frozen at -80° C in FBS + 10% DMSO before being thawed and acquired on a CyTOF Helios at either the Columbia University HIMC or the Mayo Clinic Immune Monitoring Core (IMC). All samples were acquired with, and data was normalized using spiked in four-element EQ Calibration Beads (Fluidigm/Standard BioTools) for longitudinal and cross-platform standardization.

### Single Cell RNA Sequencing and Analysis

Cells were processed for scRNAseq using the 10X Chromium 3’ v2 Library and Gel Bead Kit (10x Genomics) according to manufacturer’s instructions at the Columbia University HIMC and as described previously^26^. ScRNASeq data were processed with Cell Ranger software at the Columbia University Single Cell Analysis Core. Illumina base call files were converted to FASTQ files with the command “cellranger mkfastq.” Expression data were processed with “cellranger count” on the pre-built human reference set of 30,727 genes. Cell Ranger performed default filtering for quality control. The Seurat package was used for quality control filtering of cells, excluding those with fewer than 10% mitochondrial RNA content, more than 1,500 unique UMI counts, and fewer than 15,000 unique UMI counts. Pooled distribution across all samples of UMI counts, unique gene counts, and percentage of mitochondrial DNA after QC-filtering is shown in Supplementary Figure 3A.

*Gene Expression Clustering and Initial Cell Type Inference |* Combined and normalized gene expression matrix was visualized by UMAP projection and unsupervised clustering was performed using the Louvain algorithm applied to top50 principal components of the full data matrix, with optimal clustering resolution determined by bootstrapped silhouette score optimization, as described in prior work^26^. Broad cell types were inferred by SingleR with blueprint-ENCODE reference of sorted cell types, with each cluster consensus-labelled by majority-inferred cell type^28^. Further re-labelling of tumor cell clusters (labelled as epithelial by SingleR) was performed by InferCNV analysis, demonstrating consistent regions of chromosomal aberration above background rate seen in other cell types^30^.

*Regulatory Network Inference |* For each gene expression cluster, a gene regulatory network was inferred on log-normalized counts by the ARACNe algorithm^55^ to identify downstream transcriptional targets of regulatory proteins. ARACNe was run with 100 bootstrap iterations using 1785 transcription factors (genes annotated in gene ontology molecular function database as GO:0003700, “transcription factor activity,” or as GO:0003677, “DNA binding” and GO:0030528, “transcription regulator activity,” or as GO:0003677 and GO:0045449, “regulation of transcription”), 668 transcriptional cofactors (a manually curated list, not overlapping with the transcription factor list, built upon genes annotated as GO:0003712, “transcription cofactor activity,” or GO:0030528 or GO:0045449), 3455 signaling pathway related genes (annotated in GO biological process database as GO:0007165, “signal transduction” and in GO cellular component database as GO:0005622, “intracellular” or GO:0005886, “plasma membrane”), and 3620 surface markers (annotated as GO:0005886 or as GO:0009986, “cell surface”).

ARACNe is only run on these gene sets so as to limit protein activity inference to proteins with biologically meaningful downstream regulatory targets, and we do not apply ARACNe to infer regulatory networks for proteins with no known signaling or transcriptional activity for which protein activity may be difficult to biologically interpret. Parameters were set to zero DPI (Data Processing Inequality) tolerance and MI (Mutual Information) p value threshold of 10−8, computed by permuting the original dataset as a null model.

*Protein Activity Inference and Clustering |* Inference of protein activity was performed from scaled integrated gene expression matrix and the set of all ARACNe-inferred gene regulatory networks using the VIPER algorithm^24,26^. Normalized Enrichment Scores of protein activity were successfully inferred on a per-cell basis for 3,437 regulatory proteins. This protein activity matrix was re-clustered in Seurat using the Louvain algorithm on top50 principal components and clustering resolution parameter optimized by silhouette score, as above, with two-dimensional data projection visualized by UMAP. Differential protein activity between clusters was computed by Wilcoxon rank-sum test with multiple testing correction. Differential abundance of each cluster per patient by treatment group was also assessed by Wilcoxon-rank-sum test, as well as differential abundance of each cluster within the combination-treatment group between patients with and without disease recurrence. Cell clusters inferred by SingleR as lymphoid lineage, myeloid lineage, or tumor cells, respectively, were each isolated for further sub-clustering analyses.

*Single-cell RNA-seq T/NK Subclustering |* Subsetting lymphoid cells from initial analysis resulted in a VIPER matrix of 14,208 cells, projected into their first 50 principal components using Seurat’s RunPCA function, followed by using Seurat’s RunUMAP function with method umap-learn. Unsupervised clustering of lymphoid cells was performed by the same silhouette score optimized Louvain approach as above. In addition to SingleR cell type labelling, we applied GSEA on cell-by-cell basis using an expression-based T-reg marker gene set previously published from Obradovic et. al. Cell 2021^26^, to identify the T-reg cluster. VIPER activity of manually selected relevant markers was also visualized for lymphoid cell subtype identification of all other clusters. Differential abundance of each cluster as percent of lymphoid cells was assessed by treatment group and by recurrence vs non-recurrence within the combination-treatment group, by Wilcox rank-sum test. The ADT-only treatment group was not stratified due to insufficient recurrences for statistical power and recurrence data were not available in the untreated group. Further testing in the combination treatment group of association between Treg abundance and time-to-recurrence was performed by Kaplan-Meier analysis, with optimal cut-point of high-Treg vs low-Treg samples determined by log-rank maximization, and p-value assessed by Cox regression.

*Single-cell RNA-seq Myeloid Cell Subclustering |* Isolation of myeloid cells (clusters identified by SingleR as predominantly Monocyte or Macrophage) yielded 5771 Myeloid cells, subclustered in the same manner as lymphoid cells above. RunPCA and RunUMAP from Seurat were used to compute the first 50 principal components and UMAP respectively. Upon sub-clustering, a subpopulation identified as fibroblasts (n=183) by SingleR with the BlueprintEncode reference dataset from Celldex was removed from downstream analysis. A 7-cluster unsupervised clustering solution was selected as optimal by maximization of silhouette score. Clusters were then annotated by cell type with a combination of visualizing manually selected phenotypic markers and assessing Gene Set Enrichment of differentially active proteins by cluster.

*Correlation Between FCGR3A Activity and Treg Frequency |* In order to assess for relationship between FCGR3A protein activity among myeloid cells and Treg frequency, we performed correlative analysis between Treg frequency as a percent of lymphoid cells for each patient against average FCGR3A protein activity for myeloid cells as a whole in the same patient, as well as for the specific subset of TREM2+ TAMs.

Stratifying these analyses by treatment group shows treatment-dependent association between FCGR3A activity and Treg abundance, such that in combination-treatment alone higher myeloid FCGR3A activity was found to associate with lower Treg abundance.

*Single-cell RNA-seq CD8+ T Cell Subclustering |* Upon sub-clustering lymphoid cells, we further isolated the two identified clusters of CD8 T-cells (CD8-1 and CD8-2) for deeper sub-phenotyping. This yielded a VIPER protein activity matrix for 7,475 cells. Re-clustering on the top50 principal components of this matrix yielded a four-cluster solution such that one small subset of n=187 cells appeared to be naïve CD8s or potential contamination and were removed. This conclusion was supported by GSEA on the gene expression signatures of these cells with the Naive CD8 gene set from Zheng et al. 2021^38^. The remaining true CD8+ cells were re-scaled and re-clustered yielding a three-cluster solution. These clusters were labelled as “Central Memory”, “Effector” and “Effector/Resident Memory” by Gene Set Enrichment Analysis. “Memory vs Effector” and “Effector vs Memory” signatures were generated using a Mann-Whitney U-Test between the gene expression profiles of CD8+ Trm and CD8+ cytotoxic populations reported by Tuong et al. 2021^34^. The top genes from each of these signatures and in the signatures from Zheng et al. 2021^38^ were supplied as gene sets to GSEA to compute enrichment on the gene expression profiles of our CD8+ cells. Relevant manually curated marker genes were further visualized by VIPER activity per cluster. Cluster frequencies by treatment group were then calculated to identify changes in these CD8+ subtypes in response to treatment.

### Immunofluorescence

Archival FFPE prostate tumor specimens were selected by a board-certified pathologist. Immunoblank slides and representative adjacent H&E slides were sectioned at 5uM by the Herbert Irving Comprehensive Cancer Center’s Molecular Pathology Shared Research (HICCC MPSR). Tissue sections were deparaffinized using a standard series of xylene and ethanol washes and rinsed with distilled water. Antigen retrieval was performed by heating the slides in citrate buffer at pH of 6.0 for 20 minutes in a pressure cooker. Slides were incubated overnight at 4°C with primary antibodies against CD4 (1:200, Abcam) and FOXP3 (1:100, Invitrogen). Subsequently, the slides were incubated for 30 minutes at room temperature with horse anti-mouse antibody (1:200, Vector Laboratories), and then for 30 minutes with Streptavidin, Alexa Fluor™ 594 conjugate (1:1000, Invitrogen). Next, the slides were incubated for 30 minutes at room temperature with goat anti-rabbit antibody (1:200, Vector Laboratories) and then for an additional 30 minutes with Streptavidin, Alexa Fluor™ 488 conjugate (1:300, Invitrogen). Slide sections were mounted at room temperature with 25uL of VECTASHIELD Vibrance® Antifade Mounting Medium with DAPI (Vector Laboratories) and coverslipped.

Fluorescent images were acquired by the PhenoImager™ HT Instrument (Akoya Biosciences). Onboard spectral unmixing was performed using the PhenoImager HT 2.0 software (Akoya Biosciences). The images were analyzed with QuPath version 0.5.1^56^. Regions of interest were generated referencing tumor-marked H&E slides from adjacent sections. To further refine tumor region selection, ROI annotation was guided by predictions from the prostate-tumor-resnet34.tcga-prad model in the WSInfer QuPath extension— a deep learning model trained on TCGA prostate whole slide images to differentiate tumor from benign tissue^57^. Nuclear and cell segmentation were performed using the QuPath extension StarDist 0.5.0 with the pretrained model dsb2018_heavy_augment.pb^58^. Following cellular segmentation, object classifiers were trained on each image to quantify Tregs (CD4+/FOXP3+) per mm^2^ or as a percentage of all DAPI+ cells.

### Mice

Male FVB mice were purchased from Jackson Laboratory (Bar Harbor, ME). Mice were 8-10 weeks old at time of use. All animals were housed in strict accordance with NIH and American Association of Laboratory Animal Care regulations. All experiments and procedures for this study were approved by the Mayo Clinic Institutional Animal Care and Use Committee (IACUC).

### Animal Studies

MycCaP cells were provided as a kind gift from Dr. Charles Drake and were verified as free of pathogen and cell line contamination using a 19-marker mouse STR panel through IDEXX BioAnalytics (Columbia, MO). MycCaP cells were passaged in RPMI medium (Corning; Corning, NY) supplemented with 10% FBS (Corning; Corning, NY), 100 U/mL penicillin, and 100 mg/mL streptomycin (Gibco; Gaithersburg, MD). For tumor implantation, Myc-CaP cells at 70-90% confluence were harvested with 0.05% trypsin (Corning; Corning, NY), washed with PBS, counted, and resuspended at 10x10^6^ cells/mL in ice cold PBS. On day 0, 8–10-week-old mice were implanted on the right flank with 1x10^6^ MycCaP cells. Cages were randomized to treatment groups after cages were manually normalized to reduce intergroup variability, using the final tumor volume measurement values available prior to treatment. Tumor measurements were recorded in X, Y, and Z dimensions in a non-blinded fashion (largest diameter, smallest diameter, and largest height, respectively) every 2-3 days by digital caliper and tumor volume was calculated by multiplying X*Y*Z. On day 24 when tumors were approximately 200mm^3^ on average, mice received a single subcutaneous injection in the neck scruff of 625 ug degarelix acetate (ADT; Cayman Chemical; Ann Arbor, MI) resuspended in 100 μl sterile water per mouse. Appropriate treatment groups additionally received anti-CTLA4 (D) (9D9-mIgG2a) or anti-CTLA4 (ND) (9D9-mIgG2a-LALAPG) as indicated by intraperitoneal (IP) injection at 100 μg per mouse in 100 μl sterile PBS on days 24, 27, and 30. Antibodies were purchased from BioXCell (Lebanon, NH). For flow cytometry analysis, cohorts of animals were euthanized on day 32 for tissue collection and processing. For survival analysis, mice were deemed eligible for euthanasia when tumor volume exceeded 1,000 mm^3^ or tumor ulceration exceeded 5 mm in diameter.

### Spectral Flow Cytometry

*Tissue harvest and dissociation |* Following mouse euthanasia, tumor-draining inguinal lymph nodes were dissected and placed in 48-well plates containing 150 μl R10 media on ice. Tumors were harvested, massed, and up to 50 mg tumor was diced and placed in X-Vivo 15 media (Lonza; Basel, Switzerland) in 15mL Eppendorf tubes on ice. DNase (40 μl of 20 mg/mL solution; Roche; Basel, Switzerland) and Collagenase D (125 μl of 40 mg/mL solution; Roche; Basel, Switzerland) were added to tumor samples prior to incubation on a shaker at 37° C for 30 minutes. Digest reactions were quenched by adding 5mL R10 media. Tumor digests were filtered through 70 μm filters (Miltenyi; Bergisch Gladbach, GE), remaining undigested tissue was mashed with the handle end of a 3mL syringe plunger, then filters were washed with 5mL R10 prior to centrifugation.

Lymph nodes were physically disaggregated with a 1mL syringe plunger in the 48-well plate. All samples were then transferred in R10 into U-bottom 96-well plates for staining.

*Staining protocol |* Samples were washed with PBS. Dead cells were stained by resuspension in 100 μl PBS + Live/Dead Fixable Blue dye (1:500; Invitrogen; Waltham, MA). This and all further staining or fixation steps were performed for 30 minutes at room temperature on a plate shaker protected from light, unless specified otherwise.

Samples were washed 2x with PBS, then were resuspended in FACS (PBS + 3% FBS + 1mM EDTA + 10mM HEPES) supplemented with TruStain FcX (1:50; BioLegend; San Diego, CA) and TruStain Monocyte Blocker (1:20; BioLegend; San Diego, CA) and placed on ice. Surface antibodies were prepared at optimal dilutions (see Supplementary Table S1) in FACS supplemented with Brilliant Stain Plus buffer (BD; Franklin Lakes, NJ), then were added to samples in blocking solution. After staining, samples were washed 2x with FACS buffer and fixed in 100 μl of FoxP3 Fixation/Permeabilization Kit buffer (eBioscience; San Diego, CA). Samples were washed twice with 1X Permeabilization buffer (1X PW) then were stained in 1X PW plus intracellular antibodies. Plate(s) were sealed and intracellular staining was performed overnight at 4° C on an orbital shaker. Samples were then washed twice in FACS then resuspended in 200 μl FACS and were immediately acquired on a Cytek Aurora 5-laser (UV/V/B/YG/R) cytometer. Simultaneously stained splenocyte samples or SpectraComp beads (Slingshot Biosciences; Emeryville, CA) were utilized for single stain controls as indicated in Supplemental Table S1.

### Analysis of Mass Cytometry and Flow Cytometry Data

All analysis was performed using FlowJo (BD; San Diego, CA). Live, single cell, CD45+ events from all files were identified by manual gating using conventional gating strategies for CyTOF and flow cytometry data as shown in Supplementary Fig 5-6.

Samples with fewer than 1,000 live CD45+ single cell events were excluded from further analysis. All live CD45+ single cell events in the CyTOF data were concatenated for high dimensional analyses, while up to 50,000 live CD45+ single cell events from the flow cytometry dataset were downsampled and concatenated for downstream analysis. Canonical immune lineages in both datasets were defined in a semi-supervised manner utilizing FlowSOM^31^. Data were purposely overclustered to 30 FlowSOM-derived clusters based on expression of lineage-defining markers in each panel. These clusters were then manually collated based on lineage marker expression to obtain the minimal possible number of canonical immune lineages. Clusters were embedded in two-dimensional space for visualization using the supervised UMAP function in FlowJo, which performs dimensionality reduction based on selected lineage-defining marker expression with incorporation of FlowSOM-derived clusters as a weighted parameter.

For discovery-based subclustering of individual lineages (e.g. CD4 Tregs), an additional round of semi-supervised FlowSOM was performed in an analogous manner as described above; involving FlowSOM-derived overclustering based on phenotypic markers differentially expressed by cells within the lineage of interest, followed by manual collation of clusters into minimal number of phenotypically distinct subclusters. For visualization of discovery-based subclustering within lineages, we utilized PaCMAP^59^ dimensionality reduction to embed data in two dimensions. Absolute cell numbers and mean geometric fluorescent intensity (gMFI) data was exported for each cluster for downstream analysis. Phenotypic marker data was scaled based on the minimum and maximum gMFI values of each given marker across all live CD45+ cells or within the lineage of interest, as indicated in the figure legends.

### Statistical Analyses

Baseline patient characteristics and study outcomes were computed and visualized utilizing the gtsummary package (v 1.7.2)^60^, in Rstudio (R version 4.3.1, 2023-06-16). Mutational profiles were visualized using Oviz-Bio’s LandScape (v1.1.1)^61^. Stratification for Kaplain Meier analyses was performed using log-rank maximization algorithm as implemented in the survminer package. CyTOF and spectral flow cytometry statistical testing was conducted in GraphPad Prism (v10.2). All other quantitative and statistical analyses were performed using the R computational environment and packages described above. Differential gene expression was assessed at the single-cell level by the MAST single-cell statistical framework as implemented in Seurat v4^62^, and differential VIPER activity was assessed by Wilcoxon rank-sum test, each with Benjamini-Hochberg multiple-testing correction. Unless otherwise noted, measurements were taken from distinct samples and comparisons of cell frequencies by treatment group were performed by two-tailed Welch’s t-test or non-parametric Wilcox rank-sum test, and survival analyses were performed by log-rank test. In all cases, statistical significance was defined as an adjusted p value less than 0.05. Details of all statistical tests used can be found in the corresponding figure legends.

## Data Availability Statement

All data will be made available through public data repositories by time of publication.

## Code Availability Statement

All code utilized in this study is publicly available through GitHub and through utilization of published packages referenced in the study.

## Inclusion & Ethics

This clinical trial and related animal work was approved by institutional regulatory committees that involve local partners to ensure research is representative of and benefits local partners while avoiding any potential risks to the health, safety, or reputation of said partners.

## Acknowledgments

We would like to acknowledge the Columbia University Human Immune Monitoring Core (HIMC) for assistance with single cell RNA sequencing and the Mayo Clinic Immune Monitoring Core for assistance with CyTOF data acquisition. We would also like to acknowledge our collaborators at Bristol Myers Squibb for their help in conducting the study. Most of all we thank the patients and their families for participating in this study.

## Supplementary Figure Legends

**Supplementary Figure 1:**
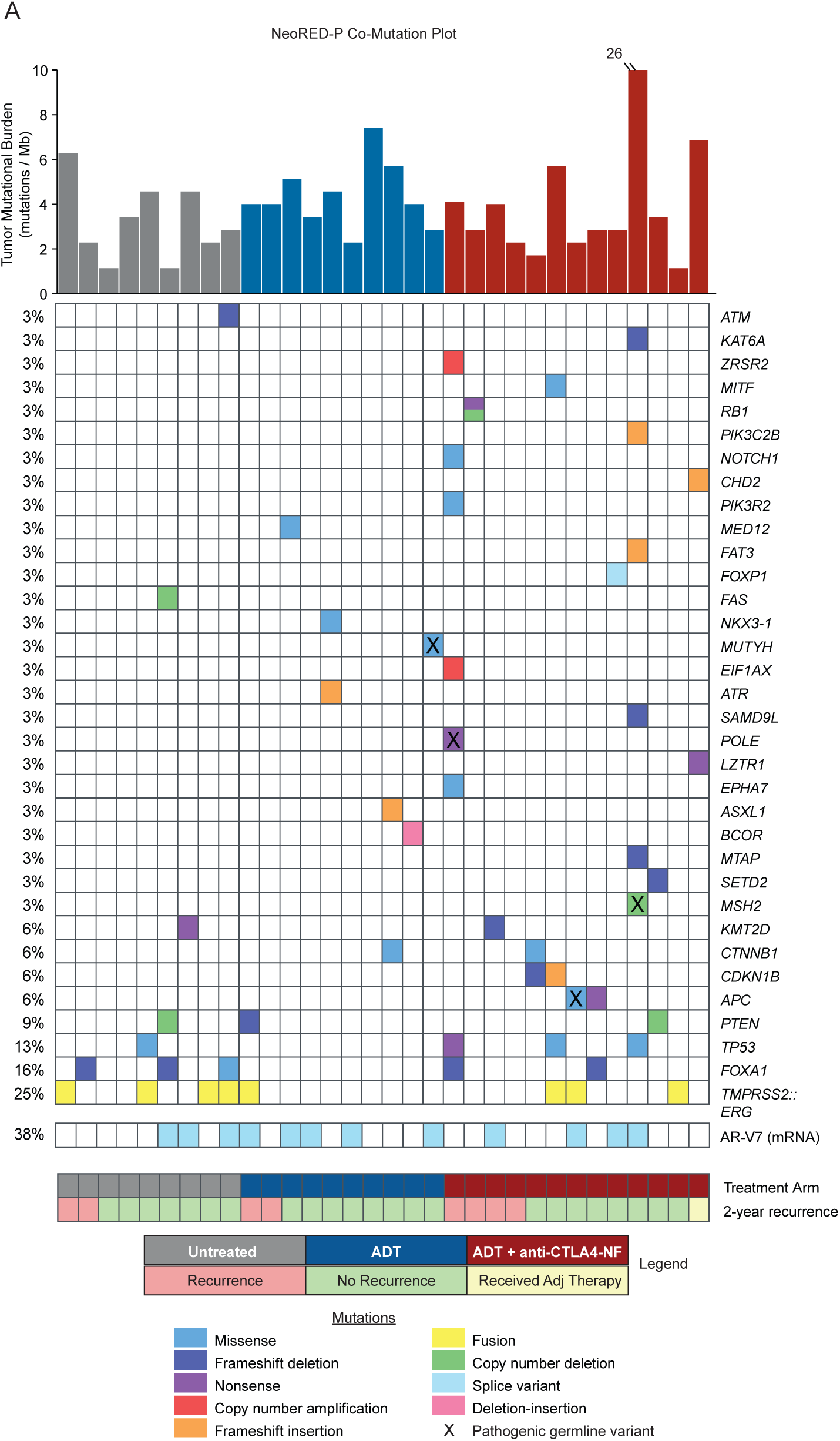
NeoRED-P study co-mutation plot. **(A)** Co-mutation plot of NeoRED-P patients and stage matched untreated control cohort.

**Supplementary Figure 2:**
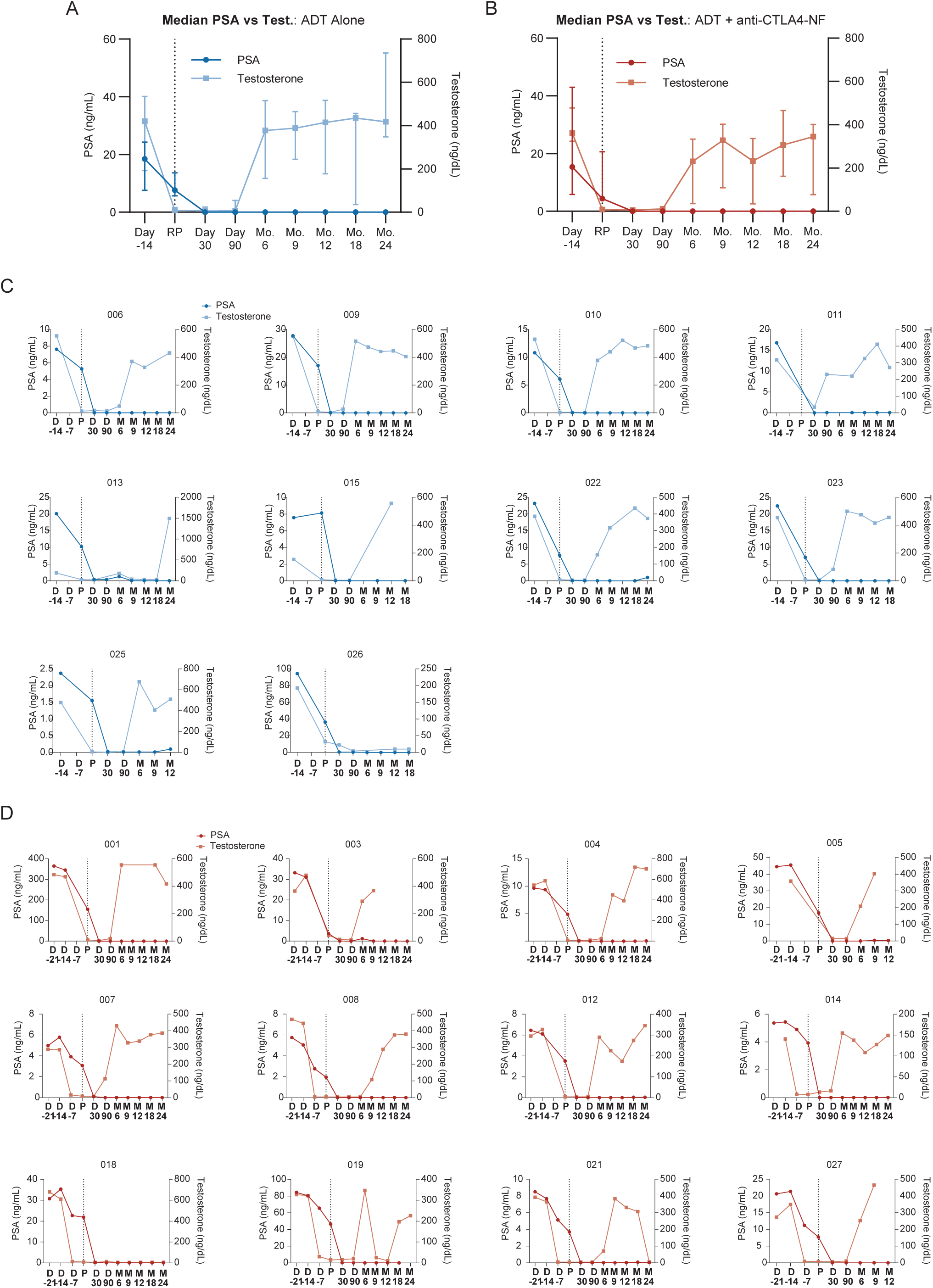
PSA responses and testosterone recovery of all trial participants. **(A)** Median serum PSA (dark blue line; round dots) and testosterone (light blue line; squares) concentrations over time of all patients treated with ADT only. Error bars denote interquartile range. **(B)** Median serum PSA (dark red line; round dots) and testosterone (light red line; squares) concentrations of all patients treated with ADT + anti-CTLA4-NF. Error bars denote interquartile range. **(C)** Plots of serum PSA and testosterone concentrations for individual patients treated with ADT only. **(D)** Plots of serum PSA and testosterone concentrations for individual patients treated with ADT + anti-CTLA4-NF. D = day; P or RP = radical prostatectomy; M or Mo = month.

**Supplementary Figure 3:**
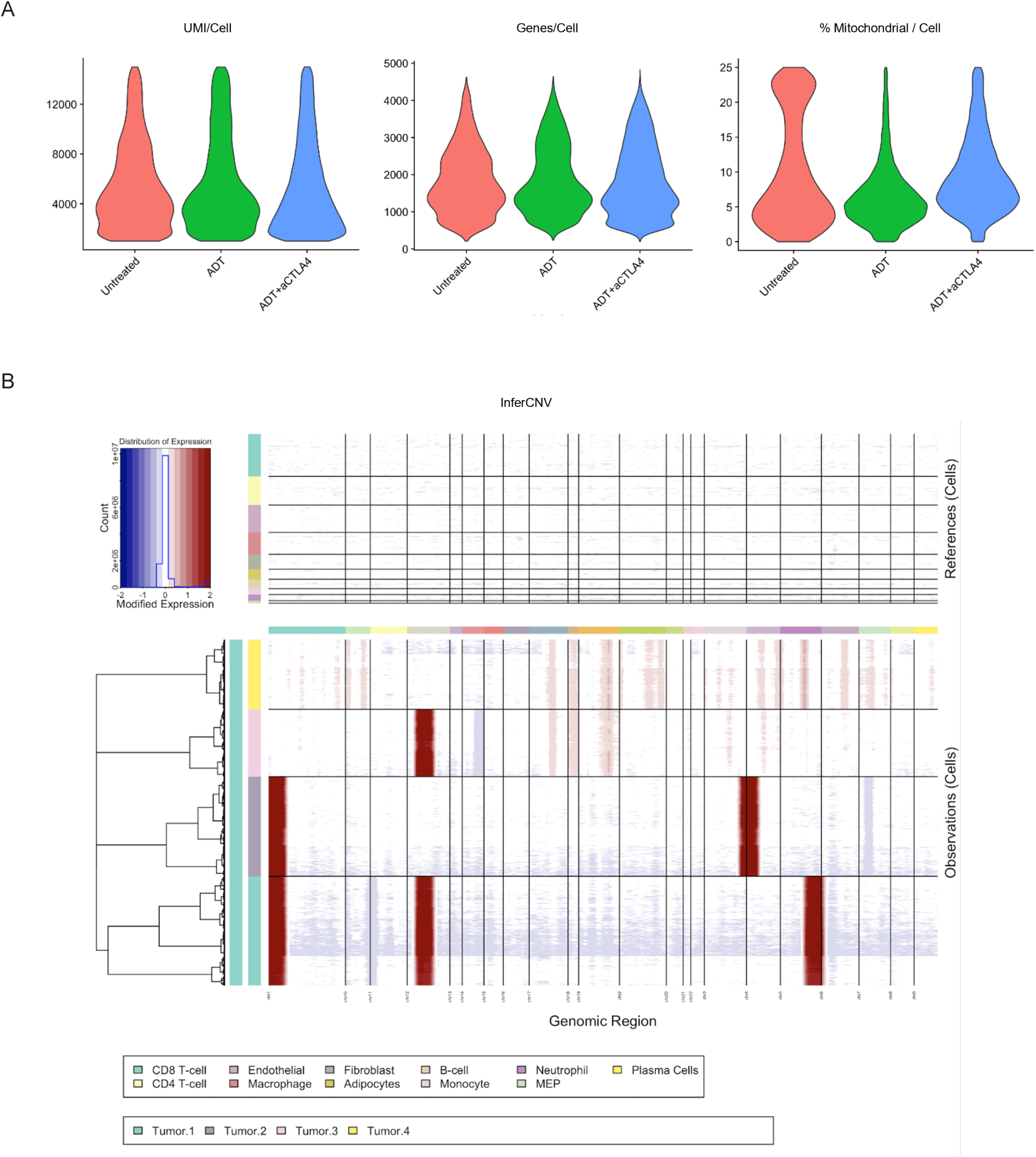
Single cell RNA sequencing QC and inferCNV results. **(A)** Violin plots representing quality control plots of scRNAseq data stratified by treatment group, including number of unique molecular identifiers (UMIs) per cell (left), number of unique genes detected per cell (middle), and percent mitochondrial gene content per cell (right). **(B)** Plots representing outcomes of InferCNV analysis through visualization of inferred copy number aberrations. Reference immune and stromal cell populations shown above, and clusters annotated as tumor cells shown at bottom.

**Supplementary Figure 4:**
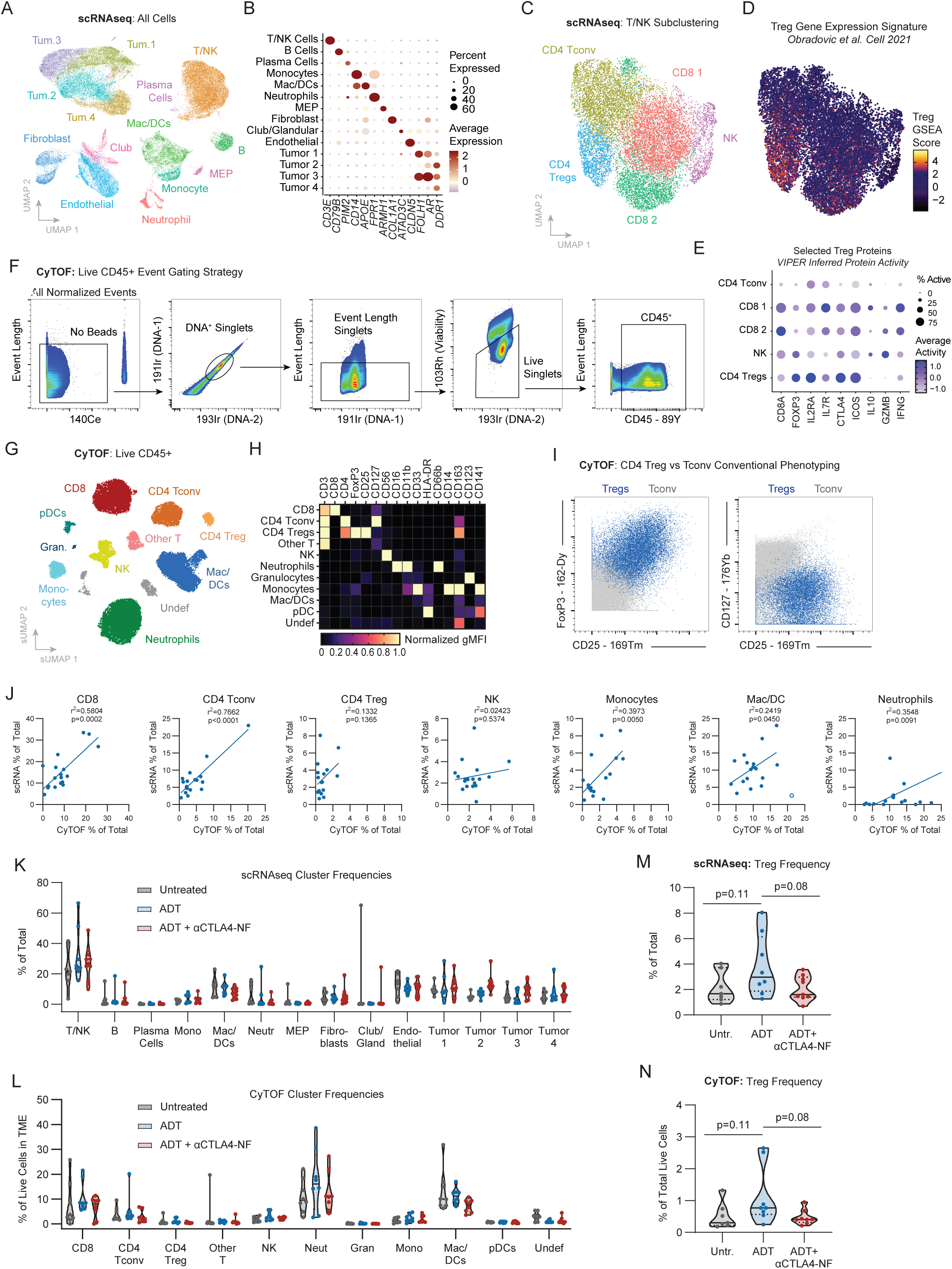
Extended NeoRED tumor scRNAseq and CyTOF data analysis. **(A)** UMAP plot of single cell RNAseq data from all patients, clustered using the PICSES analytical pipeline. **(B)** Bubble plot of representative gene expression for each cluster identified in **(A)**. **(C)** UMAP plot representing subclustering of the scRNAseq T/NK cluster shown in **(A)**. **(D)** Visualization of per-cell normalized enrichment scores for a published Treg gene set across all T/NK cells in the scRNAseq dataset. **(E)** Bubble plot of inferred protein activity for manually selected Treg-related proteins across all T/NK subclusters as calculated by VIPER. **(F)** Representative gating strategy representing isolation of viable, single cell CD45+ cells in the CyTOF dataset. **(G)** Supervised UMAP plot of CyTOF data from all patients, clustered using FlowSOM. **(H)** Heatmap of lineage-defining protein expression across all clusters. Data represents geometric MFI for each marker normalized to the minimum and maximum values across all clusters. **(I)** Overlay of FlowSOM-derived Treg and CD4 Tconv clusters in the CyTOF data on bi-axial plots representing canonical Treg phenotypes. **(J)** Pearson correlation plots comparing frequency of immune lineages annotated during clustering of both CyTOF and scRNAseq datasets as a percent of total cells. **(K)** Violin plots representing frequency and distribution of all lineage frequencies as percent of total cells by scRNAseq, stratified by treatment group. **(L)** Violin plots representing frequency and distribution of all lineage frequencies as percent of total cells by CyTOF, stratified by treatment group. **(M)** Violin plot representing Treg frequencies as a percent of total cells iterated by treatment group in the scRNAseq data. Untreated n=7; ADT n=8; ADT + anti-CTLA4-NF n=9. **(N)** Violin plot representing Treg frequencies as a percent of total cells iterated by treatment groups in the CyTOF data. Untreated n=8; ADT n=8; ADT + anti-CTLA4-NF n=8.

**Supplementary Figure 5:**
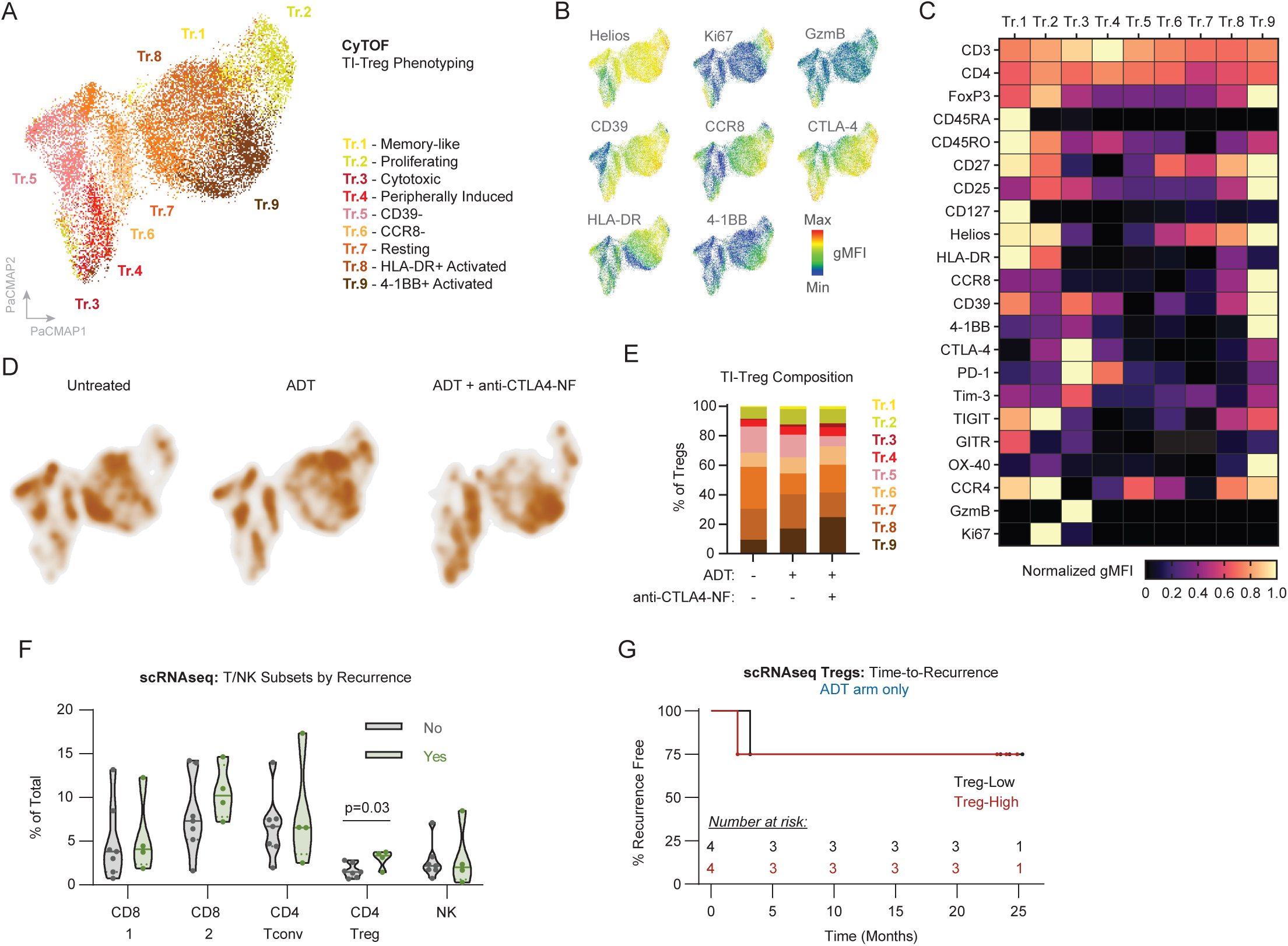
Extended phenotypic analysis of TI-Tregs and associations with clinical outcomes. **(A)** PaCMAP plot representing FlowSOM-derived Treg clusters in the CyTOF data. Data presents 17,757 total cells across 25 patients with evaluable data. Clusters were manually annotated according to expression of relevant protein markers as visualized in **(B)** as colorimetric expression maps of selected markers overlayed on the parent PaCMAP plot, or **(C)** heatmap expression of all relevant markers across all clusters. Data represents geometric MFI for each marker normalized to the minimum and maximum values across all clusters. Markers with universally high or low expression were scaled according to the minimum and maximum of all CD45^+^ FlowSOM clusters. **(D)** Density plots representing Treg cell location on the PaCMAP plot in **(A)** iterated by treatment group. **(E)** Stacked box plot quantifying relative frequencies of all Treg FlowSOM-derived clusters as a percent of Tregs the PaCMAP plot in **(A)** iterated by treatment group. **(F)** Violin plot representing frequency of T/NK subclusters in the scRNAseq data as a percent of all cells, stratified by post-surgical 2-year recurrence status. Welch’s t-test was used to evaluate statistical significance. **(G)** Kaplan-Meier curve representing time-to-PSA recurrence in ADT-treated patients stratified by Treg frequency in scRNAseq data.

**Supplementary Figure 6:**
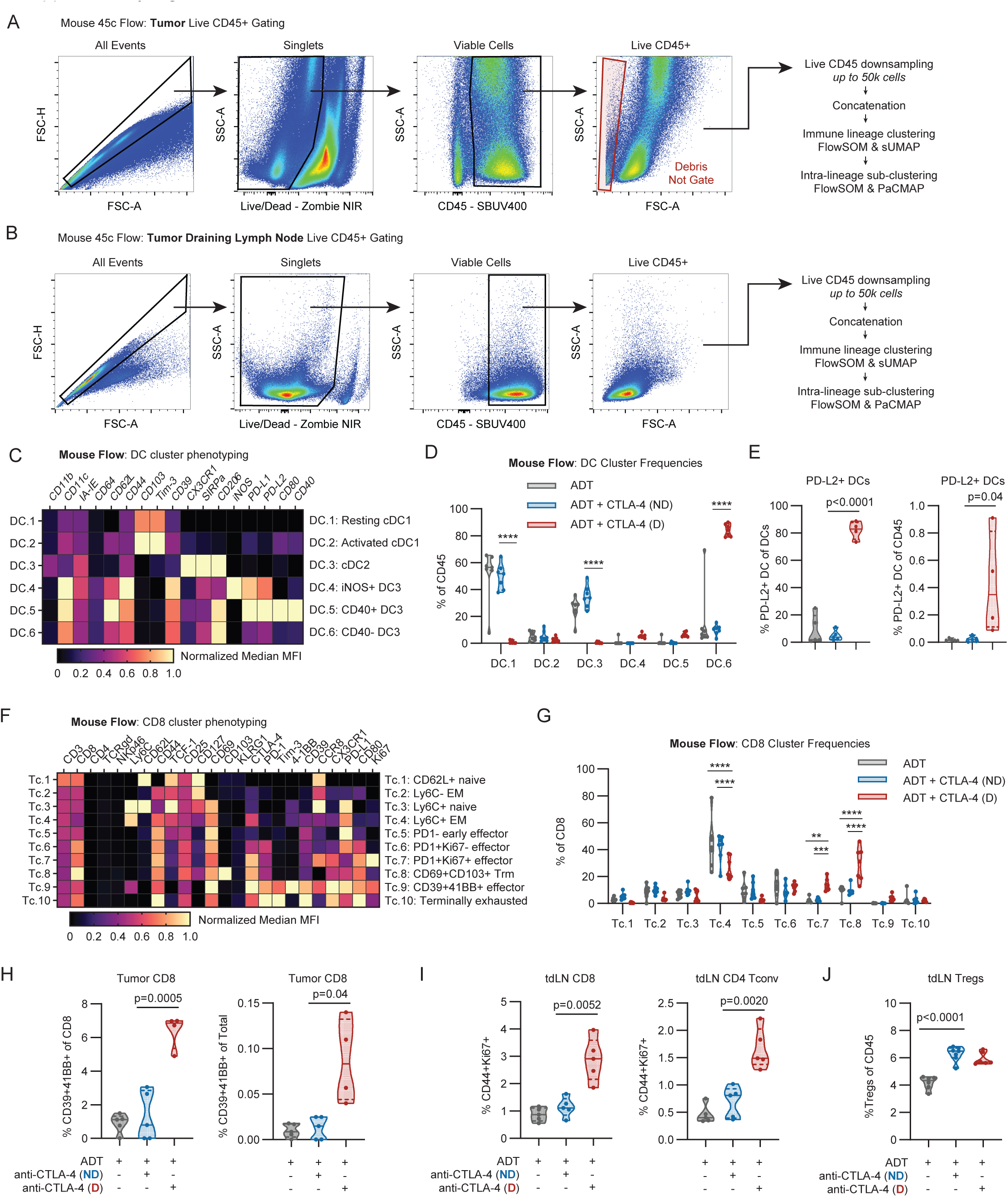
Additional flow cytometry data related to ADT + anti- CTLA4 (D) in the MycCaP model. Representative gating strategy of viable, single cell CD45+ cells in the spectral flow cytometry dataset derived from **(A)** tumor and **(B)** tumor draining lymph nodes. Summary of downstream analytical workflow shown at right. **(C)** Heatmap of relevant phenotypic marker expression across all tumor-infiltrating DC clusters. Data represents median MFI for each marker normalized to the minimum and maximum values across all DC clusters. Any markers exhibiting uniform low expression were normalized to the minimum and maximum values across all immune clusters to demonstrate low expression levels across clusters. **(D)** Violin plots representing relative proportions of DC clusters as a percentage of all DCs in the indicated treatment groups. Two-way ANOVA with Tukey multiple testing correction was used to assess statistical significance. **(E)** Violin plots representing the frequency of PD-L2+ DCs as a percentage of DCs (left) or as a percentage of CD45 (right) in a second independent replicate experiment. Student’s t-test was used to assess statistical significance. **(F)** Heatmap of relevant phenotypic marker expression across all tumor-infiltrating CD8 clusters in the mouse spectral flow cytometry data. Data represents median MFI for each marker normalized to the minimum and maximum values across all CD8 clusters. Any markers exhibiting uniform high or low expression were normalized to the minimum and maximum values across all immune clusters to demonstrate high or low expression levels across clusters, respectively. **(G)** Violin plots representing relative proportions of CD8 clusters as a percentage of all CD8s in the indicated treatment groups. Two-way ANOVA with Tukey multiple testing correction was used to assess statistical significance. **(H)** Violin plots representing the frequency of CD39+41BB+ CD8s as a percentage of tumor-infiltrating CD8s (left) or as a percentage of CD45 (right) in a second independent replicate experiment. Student’s t-test was used to assess statistical significance. **(I)** Violin plots representing the frequency of CD44^+^Ki67^+^ CD8s as a percentage of tdLN CD8s (left) or CD44^+^Ki67^+^ CD4 Tconv cells as a percentage of tdLN CD4 Tconv cells (right) in a second independent replicate experiment. **(J)** Violin plots representing the frequency of Tregs as a percentage of all tdLN CD45^+^ cells stratified by treatment group. Welch’s t-test was used to assess statistical significance.

**Supplementary Figure 7:**
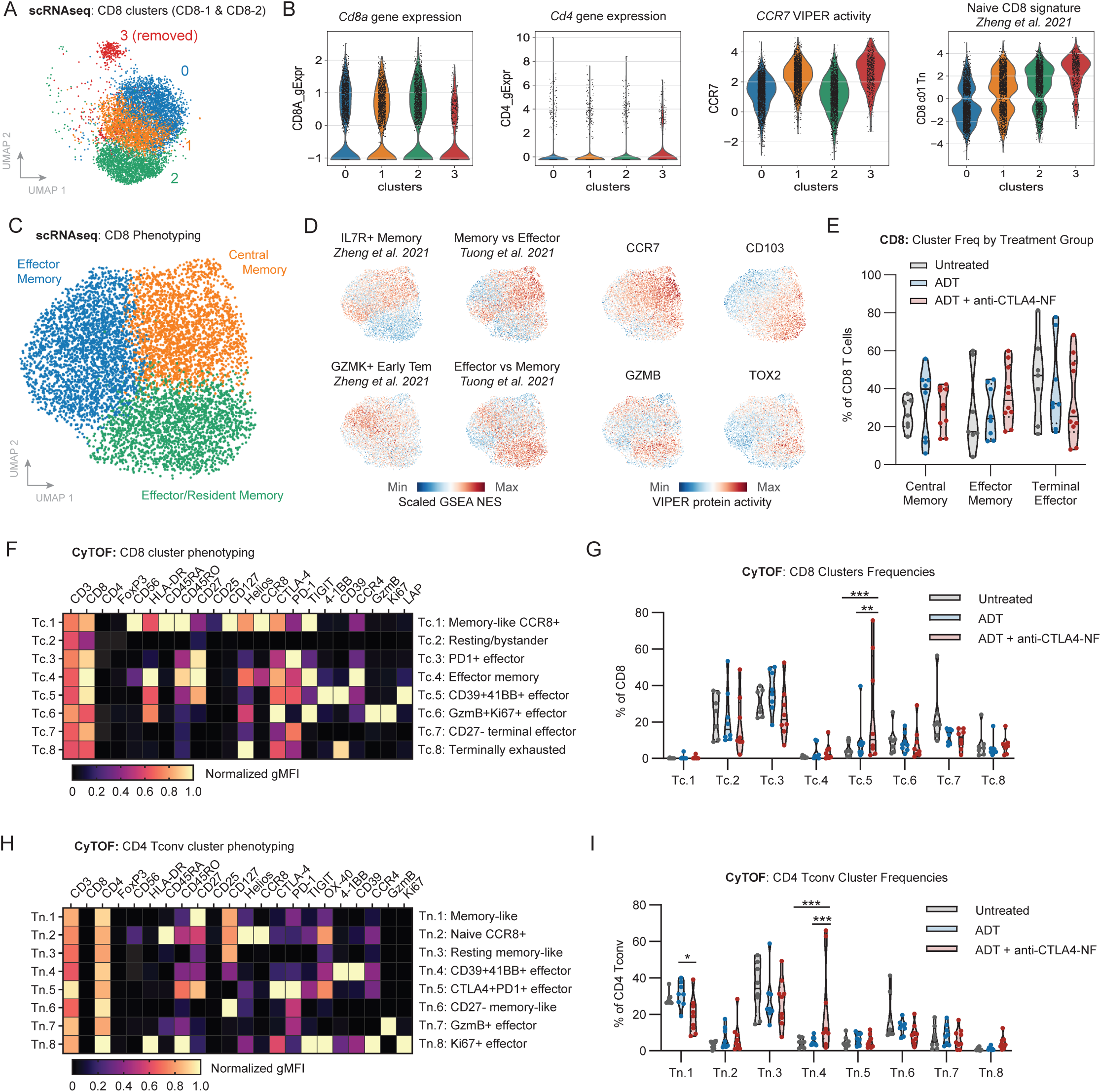
Additional T cell phenotyping data by scRNAseq, CyTOF, and flow cytometry. **(A)** UMAP plot representing subclustering of combined CD8-1 and CD8-2 clusters in the original scRNAseq T/NK cluster, identifying 4 phenotypically unique subclusters. **(B)** Violin plots representing expression of *Cd8a* and *Cd4*, VIPER inferred protein activity of *CCR7*, and tumor-infiltrating naïve CD8 T cell GSEA scores (derived from Zheng *et al*. 2021) on a per cell basis in the scRNAseq dataset, stratified by subcluster. **(C)** UMAP plot representing subclustering of CD8 T cells in the scRNAseq data utilizing the PICSES pipeline. **(D)** Visualization of representative gene expression signatures or inferred activity of indicated proteins overlayed on the parent UMAP plot to validate cluster annotations. **(E)** Violin plot of CD8 T cell cluster frequencies as a percent of CD8 T cells stratified by treatment group. Untreated n=7; ADT n=8; ADT + anti-CTLA4-NF n=10. **(F)** Heatmap of relevant phenotypic marker expression across all tumor-infiltrating CD8 clusters in the CyTOF data. Data represents geometric MFI for each marker normalized to the minimum and maximum values across all CD8 clusters. Any markers exhibiting uniform high or low expression were normalized to the minimum and maximum values across all immune clusters to demonstrate high or low expression levels across clusters, respectively. **(G)** Violin plots representing relative proportions of CD8 clusters as a percentage of all CD8s in the indicated treatment groups. Two-way ANOVA with Tukey multiple testing correction was used to assess statistical significance. **(H)** Heatmap of relevant phenotypic marker expression across all tumor-infiltrating CD4 Tconv clusters in the CyTOF data. Data represents geometric MFI for each marker normalized to the minimum and maximum values across all CD4 Tconv clusters. Any markers exhibiting uniform high or low expression were normalized to the minimum and maximum values across all immune clusters to demonstrate high or low expression levels across clusters, respectively. **(I)** Violin plots representing relative proportions of CD4 Tconv clusters as a percentage of all CD4 Tconv cells in the indicated treatment groups. Two-way ANOVA with Tukey multiple testing correction was used to assess statistical significance.

